# The congenital multiple organ malformation syndrome, Ritscher-Schinzel syndrome is an endosomal recyclinopathy

**DOI:** 10.1101/2024.08.17.24311658

**Authors:** Kohji Kato, Yosuke Nishio, Kirsty J. McMillan, Aljazi Al-Maraghi, Hester Y. Kroes, Mohamed S. Abdel-Hamid, Emma Jones, Shrestha Shaw, Aya Yoshida, Shiomi Otsuji, Yuka Murofushi, Waleed Aamer, Ajaz A. Bhat, Jehan AlRayahi, Ammira S. Alshabeeb Akil, Ellen van Binsbergen, Etienne J. Janssen, Hisashi Oishi, Ryosuke Kobayashi, Takuro Horii, Izuho Hatada, Akihiko Saito, Mitsuharu Hattori, Yoshihiko Kawano, Philip A. Lewis, Kate J. Heesom, Takeshi Takarada, Kazunobu Sawamoto, Masaki Matsushita, Tomoo Ogi, Rebeka Butkovic, Chris Danson, Kevin A. Wilkinson, Khalid A. Fakhro, Maha S. Zaki, Shinji Saitoh, Peter J Cullen

## Abstract

Ritscher-Schinzel syndrome (RSS) is a congenital malformation syndrome characterized by cerebellar, cardiac, and craniofacial malformations and phenotypes associated with liver, skeletal and kidney dysfunction. The genetic cause of RSS remains to be fully defined, and limited information is available regarding the root cause of the multiple tissue phenotypes. Here, we combine genetic and clinical analysis in patient cohorts with *in-cellulo* analysis and *in vivo* phenotypic analysis of a mouse model of RSS, to identify novel causative genes for RSS within the Commander endosomal recycling pathway. We reveal how perturbed endosomal recycling reduces tissue-specific presentation of cell surface integral membrane proteins essential for kidney, bone and brain development and how this leads to major RSS-associated clinical phenotypes including proteinuria, skeletal malformation, and neurological impairment. Our data establishes RSS as a ‘recyclinopathy’ that arises from a dysfunction in the Commander endosomal recycling pathway.

## Introduction

More than 5,500 integral membrane proteins are encoded in the human genome^1^. Different tissues display differential integral membrane protein expression, enabling each tissue and organ to perform highly specific functions. At the cell surface, integral membrane proteins including receptors, channels, transporters, and adhesion molecules play essential roles in cell communication and response to the extracellular environment, thereby controlling cell, tissue, and organism-level physiology. Their selective endocytosis and entry into the intracellular endosomal network provide an acute mechanism for modulating and adapting the functionality of the cell surface during human development and adult homeostasis^2^.

On entering the endosomal network, cell surface integral membrane proteins and associated proteins and lipids (collectively termed ‘cargo’) face a fate decision; either they are sorted to lysosome for degradation and subsequent removal from the cell, or they undergo retrieval and recycling for reuse at the cell surface^2^. Orchestrating the retrieval and recycling pathways are multiprotein complexes that include Retromer, Retriever, the CCC complex, and the WASH complex, which are present on specific retrieval subdomains on the limiting membrane of individual endosomes^3–6^. Specific cargo adaptors select cargo proteins in a sequence-dependent manner to present them to these multiprotein complexes for entry into the recycling pathway.

Retriever is a heterotrimer of VPS35L, VPS26C, and VPS29^5^, which associates with the CCC complex, a protein assembly of CCDC22 and CCDC93 and 10 COMMD proteins (COMMD1-COMMD10), and an additional protein DENND10 to form the Commander super-assembly^7–9^. Commander associates with two key accessory proteins, the ARP2/3 activating WASH complex to regulate the generation of a localised branched filamentous actin network at the retrieval subdomain^10^, and the cargo adaptor sorting nexin-17 (SNX17) that binds to integral membrane proteins containing ΦxNPxY/F or ΦxNxxY/F sorting motifs in their cytoplasmic facing domains (where Φ is a hydrophobic residue and x is any residue)^5,11–14^. Classic cargos for SNX17-mediated entry into the Retriever/CCC/WASH retrieval and recycling pathway include lipoprotein receptors and integrins^15–17^.

Loss-of-function mutations in the *VPS35L*, *CCDC22*, and *WASHC5* subunits of the SNX17-Retriever/CCC/WASH pathway are responsible for a congenital malformation syndrome, Ritscher-Schinzel syndrome (RSS)^18–25^. RSS is characterized by cerebellar, cardiac, and craniofacial malformations (hence its alternative name, 3C syndrome), but comprehensive analysis has revealed a variety of other phenotypes including lipid metabolism disorders, skeletal abnormalities, and renal disorders^18^. We, and others, have established that the observed hypercholesterolemia arises from a defect in the endosomal retrieval and recycling of hepatocyte lipoprotein receptors, LRP1 and LDLR, leading to a decrease in their cell surface expression and a reduction in the uptake of circulating LDL-cholesterol^18,26^. Both LRP1 and LDLR contain ΦxNPxY sorting motifs that are recognised by SNX17 to allow their selection for Commander-mediated retrieval and recycling^14,15^. Loss of function in this pathway leads to LRP1 and LDLR mis-sorting into the lysosomal degradative pathway resulting in reduced cell surface levels of these lipoprotein receptors^18,26^. On the other hand, DPYSL5, a member of the Dihydropyrimidinase-like (DPYSL) protein family (also termed Collapsin response mediator proteins (CRMP))^27–29^, which interacts with cytoskeletal-associated proteins such as tubulin, has been suggested to constitute a fourth gene responsible for RSS. However, as far as we are aware, there is no evidence linking DPYSL proteins to endosomal cargo retrieval and recycling. Thus, in part because of the small number of studied RSS patients, there remains a lack of knowledge regarding the root cause of RSS and the molecular mechanism leading to the clinical symptoms observed in this syndrome.

Here, we have combined genetic analysis in patient cohorts with *in vitro* and *in vivo* cellular and biochemical analyses to provide tissue specific insight into the dysregulation of the SNX17-Retriever/CCC/WASH retrieval and recycling pathway in RSS. We identify *CCDC93*, *COMMD4*, and *COMMD9*, and show *VPS26C* and *WASHC4* as novel causative genes for RSS and establish that all cases display biallelic loss of function mutations and exhibit overlapping clinical features. Targeted cellular and biochemical analyses confirm that ablation of each gene function leads to a dysregulation of SNX17-Retriever/CCC/WASH cargo retrieval and recycling, while the unbiased application of cell surface-restricted proteomics in selected tissue-types reveals the spectrum of integral membrane proteins undergoing mis-sorting and how their cell surface reduction contributes to the development of the tissue-specific clinical phenotypes in RSS. Finally, we develop a Vps35l-tissue specific knock-out mouse model to provide an *in vivo* validation of the connectivity between cellular, tissue and clinical phenotypes. Together, our findings provide new clinical and cellular insight into the contribution of SNX17-Retriever/CCC/WASH endosomal cargo retrieval and recycling in human development and human health and provides the first detailed understanding of the molecular mechanism behind many of the principal clinical phenotypes observed in RSS.

## Results

### Genetic analysis solidifies Ritscher-Schinzel syndrome is caused by defective SNX17-Retriever/CCC/WASH-mediated endosomal retrieval and recycling

The small number of patients with genetically confirmed RSS limits the confidence regarding the disease entity and the root molecular cause of this condition. Although DPYSL5, the fourth gene currently considered to be responsible for RSS, has not been linked to the endosomal recycling network, the evidence that three out of the four responsible genes (*VPS35L*, *CCDC22*, and *WASHC5*) are involved in SNX17-Retriever/CCC/WASH-mediated endosomal cargo retrieval and recycling prompted us to gather additional genetic evidence to determine whether dysregulation of this pathway is primarily attributed to RSS.

Through international collaboration fostered by GeneMatcher^30^, we identified five patients from three families, each of whom carried biallelic mutations in genes of the CCC complex, either a homozygous missense variant in *COMMD4* [NM_017828.5:c.122T>G; p.Leu41Arg (L41R)]; a homozygous frameshift variant in *COMMD9* [NM_014186.3:c.208_209del; p.Leu70GlyfsTer5 (L70Gfs*5)]; or compound heterozygous variants in *CCDC93* [NM_019044.5: c.1371T>G; p.Tyr457Ter (Y457*), c.1891T>G; p.Ser631Ala (S631A)] (Figure 1A-1C were removed due to medRxiv policy. Family trees are available upon request from the corresponding authors). All patients exhibited overlapping clinical features with previously reported RSS patients with pathogenic biallelic mutations in *WASHC5*, *CCDC22*, and *VPS35L* (Figure 1D-1U, and Table S1. Supplementary clinical information and patient photographs were removed due to medRxiv policy. These data are available upon request from the corresponding authors.)^18–23,31–33^. To define the functional significance of these mutations, we first investigated the functional alteration of the mutant proteins COMMD4-L41R and COMMD9-L70Gfs*5 by using comparative proteomics to quantify their interactomes relative to corresponding wild-type proteins. In contrast to GFP-COMMD9, GFP-COMMD9-L70Gfs*5 displayed a loss of assembly into the CCC complex and association with Retriever and DENND10 of the Commander super-assembly (Figure 1V, Figure S1A and Supplementary Data1), while COMMD4-L41R retained the ability to interact with all Commander components (Figure S1B-1D). However, COMMD4-L41R did display a significantly enhanced degradative turnover compared to wild-type COMMD4, likely reflective of a disruption in a highly conserved hydrophobic cavity by the introduction of a positively charged hydrophilic residue (Figure 1W and S1E). Consistent with decreased protein stability, HEK293T cells carrying a COMMD4-L41R knock-in exhibited a decrease in the steady-state abundance of COMMD4 (Figure 1X). Finally, Serine-631 of CCDC93 is highly conserved across species and whilst CCDC93-S631A retained the ability to form the CCC complex it also displayed a significantly enhanced degradative turnover compared to wild-type CCDC93 (Figure 1Y, Figure S1F and S1G). Consistent with enhanced protein instability, western blot analysis showed a significant reduction of CCDC93 in fibroblasts derived from the CCDC93(Y457*/S631A) patient compared to controls. Additionally, the level of CCDC22 and VPS35L expression were also significantly decreased in the patient cell line (Figure 1Z). Together, these data reveal that each identified mutation leads to a loss of function in the assembly and/or stability of the CCC complex and its association with Retriever and DENND10 within the Commander super-assembly.

**Figure 1.**
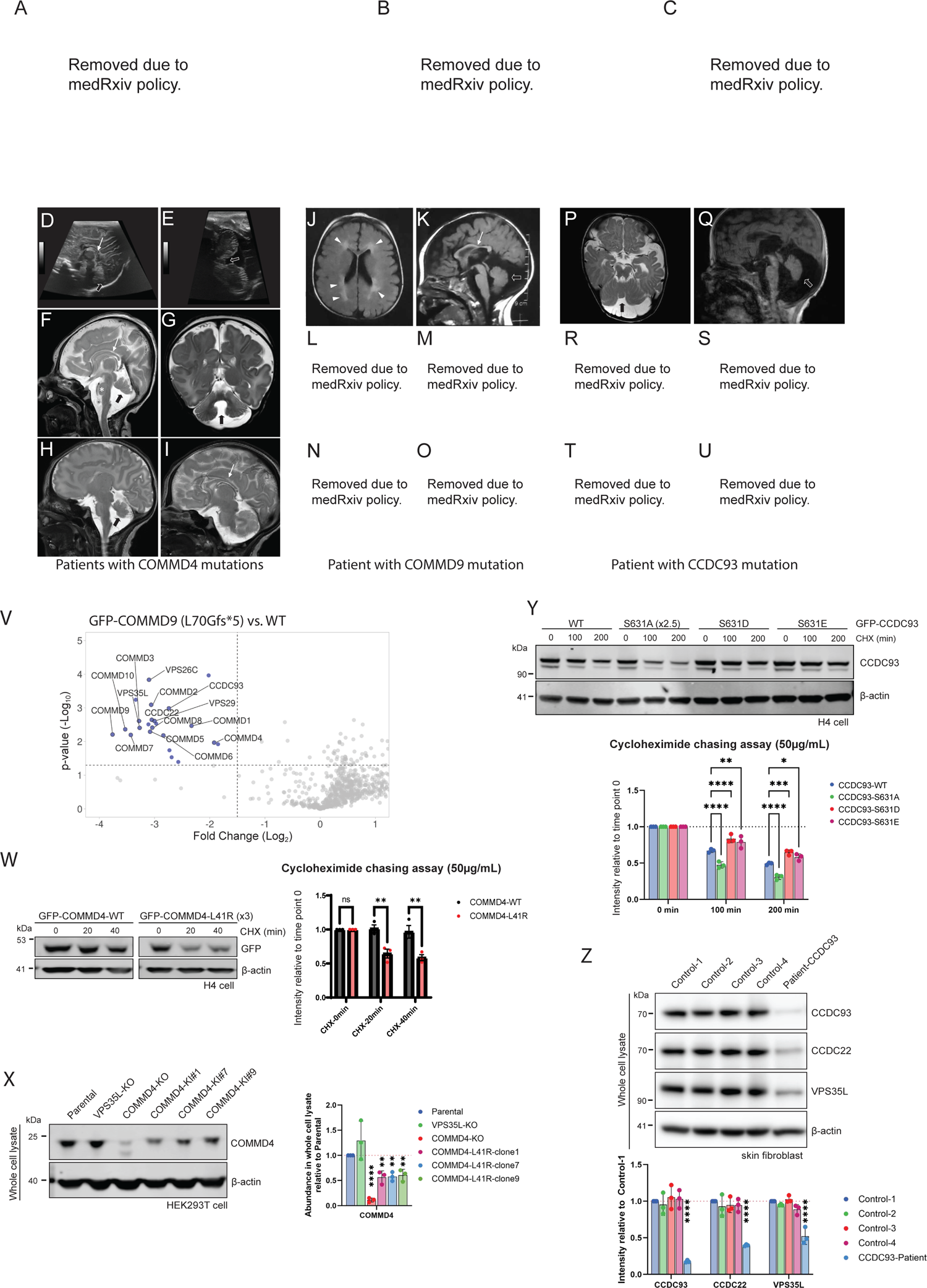
Biallelic pathogenic variants in *COMMD4*, *COMMD9*, and *CCDC93* cause Ritscher-Schinzel syndrome. (A-C) Pedigree analyses with the patients represented by a filled symbol. *COMMD4*, *COMMD9*, or *CCDC93* genotypes are given. (D-I) Brain MRI or ultrasonography of the patients with homozygous COMMD4 mutation. All show dysgenic corpus callosum (thin white arrow) and inferior cerebellar hypoplasia with cranial rotation (thick black arrow). Transcranial ultrasonography of COMMD4-II-3 was taken at ages 0-5 (D and E). T2-weighted brain MRI of COMMD4-II-4 (F and G) and COMMD4-II-5 (H and I) were taken at ages 0-5. The small pons was also observed in COMMD4-II-4 (asterisk). (J-O) Clinical features of the patient with homozygous COMMD9 mutation. (J and K) Axial slice of fluid attenuation inversion recovery image and sagittal slice of T1-weighted image taken at ages 0-5. (L-N) Facial photographs show dysmorphic features and aplasia cutis congenita on the scalp. (O) Rocker bottom and flat feet. (P-U) Clinical features of the patient with compound heterozygous CCDC93 mutations. (P and Q) Axial slice of T2-weighted image and sagittal slice of T1-weighted image taken at ages 0-5. (R) A low set, small and posteriorly rotated ear. (S) Aplasia cutis congenita on the scalp. (T and U) Short fingers in patient hands and feet. Overlapping toes and hypoplastic nails were observed. (V) Volcano plots of depleted (blue circles) interactors in GFP-COMMD9-L70Gfs*5 mutant compared to GFP-COMMD9-WT in TMT-based proteomics with three replicates. (W) Representative blots of cycloheximide (CHX) chase analysis for COMMD4-L41R mutant. H4 cells expressing GFP-COMMD4 wild-type or GFP-COMMD4-L41R were treated with CHX for the indicated period, then whole cell lysates were used for immunoblotting. Bar graph shows quantification of the band intensities determined relative to time point 0 for each experiment (n=3). To achieve comparable protein level between COMMD4-WT and COMMD4-L41R at time point 0, three times the amount of COMMD4-L41R were loaded. (X) Representative blots of HEK293T parental, VPS35L-KO, COMMD4-KO, and COMMD4-L41R knock-in cells. β-actin was used as loading control. Bar graphs show relative protein abundance (n=3). (Y) Representative blots of CHX chase analysis for CCDC93-S631A mutant. H4 cells expressing GFP-CCDC93-WT, S631A, S631D, or S631E were treated with CHX for the indicated period, then whole cell lysates were used for immunoblotting. Bar graph shows quantification of the band intensities determined relative to time point 0 for each experiment (n=3). To achieve comparable protein level between CCDC93-WT and CCDC93-S631A at time point 0, 2.5 times the amount of CCDC93-S631A was loaded. (Z) Representative blots of four controls and CCDC93 patient-derived skin fibroblast. β-actin was used as loading control. Bar graphs show relative protein abundance (n=3). (W-Z) Error bars represent mean± SD. *, P<0.05; **, P<0.01; ***, P<0.001; ****, P<0.0001.

To define the functional significance of the three novel candidate genes, *COMMD4*, *COMMD9* and *CCDC93*, we knocked out each protein as well as three known genes responsible for RSS, *VPS35L*, *CCDC22*, and *WASHC5* in H4 neuroglioma cells. Knock-out of VPS35L, CCDC22, and WASHC5 destabilized the corresponding Retriever, CCC and/or WASH complexes and resulted in the loss of endosomal localisation of the CCC complex subunit COMMD1 (Figure S2A-2C). Consistent with a loss of Commander assembly and localisation, the cell surface abundance of two classical cargo for this pathway, integrin-β1 (ITGB1) and LRP1, were significantly decreased (Figure S2D). In all cases, the knock-out of COMMD4, COMMD9, and CCDC93 showed the same phenotypes: reduced steady-state levels of VPS35L and CCDC22; loss of endosomally localised COMMD1; and a loss in the cell surface abundance of ITGB1 and LRP1 (Figure 2A and 2B). Finally, the COMMD4-L41R knock-in cells also displayed a functional defect in Commander-mediated cargo recycling, again as quantified by reduced levels of cell surface ITGB1 and LRP1 (Figure S2E).

**Figure 2.**
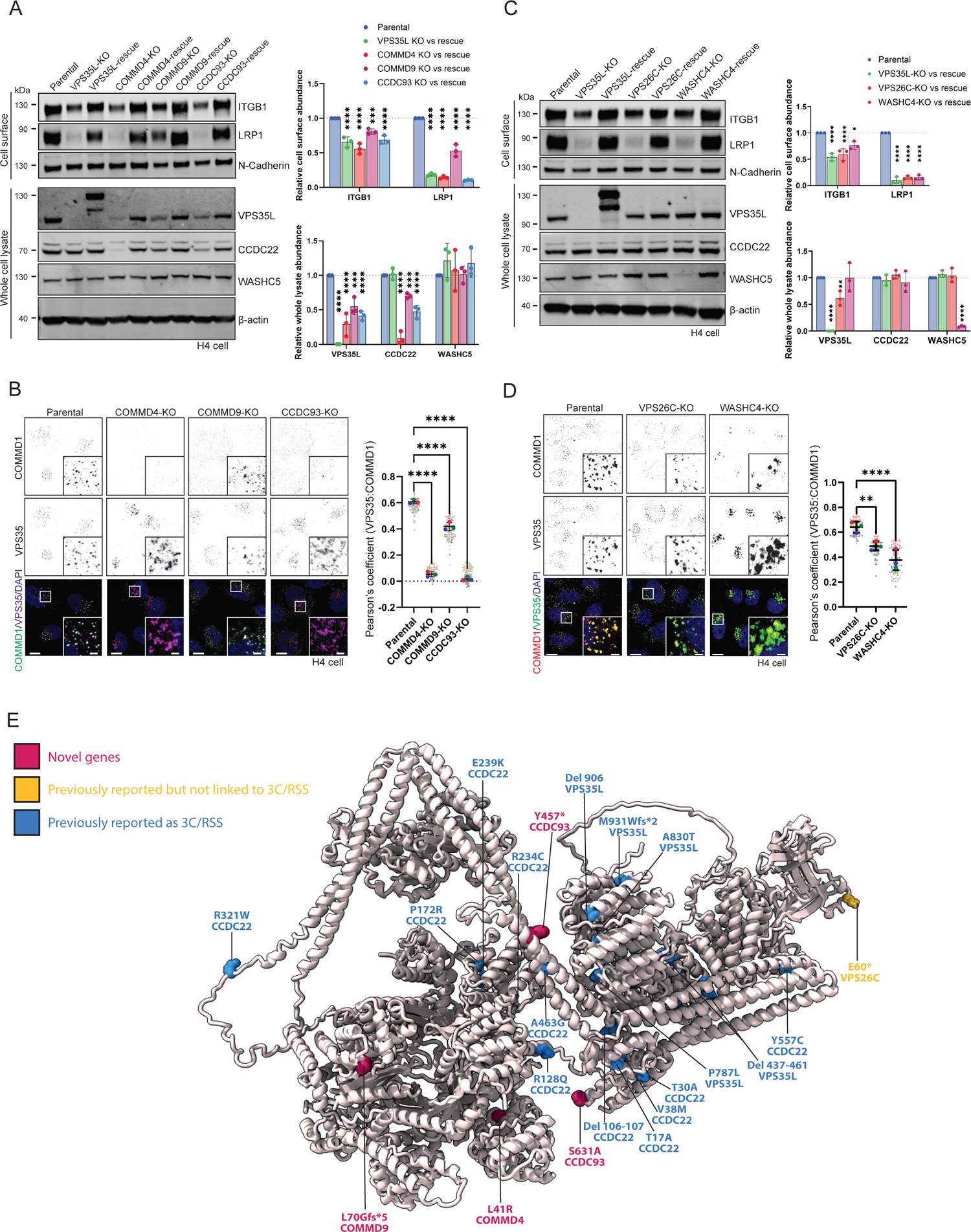
Loss of SNX17-Retriever/CCC/WASH pathway is a molecular root cause of Ritscher-Schinzel syndrome. (A) Representative blots of H4 parental and KO/rescue for VPS35L, COMMD4, COMMD9, and CCDC93. N-Cadherin and β-actin were used as loading controls. Bar graphs show relative protein abundance (n=3). (B) Representative view of immunofluorescence staining of endogenous COMMD1 and VPS35 in H4 cell lines. Scale bars, 10 µm and 2 µm, respectively. Graph shows quantification of Pearson’s correlation score from three independent experiments. Pearson’s coefficients for individual cells and means are presented by smaller and larger circles, respectively. Cell numbers analyzed across experiments were 91 parental, 86 COMMD4-KO, 93 COMMD9-KO and 90 CCDC93-KO cells. (C) Representative blots of H4 parental and KO/rescue for VPS35L, VPS26C and WASHC4. N-Cadherin and β-actin were used as loading control. Bar graphs show relative protein abundance (n=3). (D) Representative view of immunofluorescence staining of endogenous COMMD1 and VPS35 in H4 cell lines. Graph shows quantification of Pearson’s correlation score from three independent experiments. Pearson’s coefficients for individual cells and means are presented by smaller and larger circles, respectively. Cell numbers analyzed across experiments were 87 parental, 94 VPS26C-KO and 88 WASHC4-KO. (E) Mutations in VPS35L and VPS26C belonging to Retriever complex and COMMD4, COMMD9, CCDC22 and CCDC93 belonging to CCC complex associated with Ritscher-Schinzel syndrome and intellectual disability mapped onto the structure of Commander complex. (A-D) Error bars represent mean± SD. *, P<0.05; **, P<0.01; ***, P<0.001; ****, P<0.0001

Biallelic mutations in the Retriever subunit *VPS26C*, generating an early premature stop codon [NM_006052:c.178G>T; p.Glu60Ter] and the WASH complex subunit *WASHC4*, [NM_015275.2:c.1324C>T; p.Gln442Ter, c.3143A>G; p.Asp1048Gly, c.3236A>G; p.Lys1079Arg, c.1508A>G; p.His503Arg, c.3041A>G; p.Tyr1014Cys, c.3056C>G; p.PRO1019ARG], have also been observed in patients exhibiting overlapping phenotypes with RSS, although they were not clinically recognized as RSS at the time (Table S2)^34–36^. Depletion of VPS26C or WASHC4 in human H4 neuroglioma cells affected the steady-state level of VPS35L or WASHC5, suggestive of a loss in stability of the respective Retriever and WASH complexes. Consistent with this, VPS26C or WASHC4 knock-out H4 cells displayed a loss in the endosomal localisation of COMMD1 and a reduction in the cell surface abundance of ITGB1 and LRP1 (Figure 2C and 2D).

Overall, these biochemical and functional data are consistent with the identified biallelic mutations in *COMMD4*, *COMMD9*, and *CCDC93* and evidence of *VPS26C* and *WASHC4* mutations in patients with RSS-like phenotypes, being causative for RSS through a loss of function in the SNX17-Retriever/CCC/WASH endosomal cargo retrieval and recycling pathway (Figure S2F).

### An additional RSS associated gene DPYSL5 does not regulate the SNX17-Retriever/CCC/WASH endosomal retrieval and recycling pathway

A confounding factor in assigning the SNX17-Retriever/CCC/WASH recycling pathway as causative for RSS is evidence suggestive of *DPYSL5*, as a fourth gene responsible for RSS^28,37–39^. While DPYSL5 patients show some overlapping features such as cerebellar dysplasia, their clinical features do not entirely overlap with RSS (Table S2). For instance, all eight cases with the DPYSL5-E41K mutation exhibited agenesis of the corpus callosum, which has not been reported in RSS cases with mutations in genes of the SNX17-Retriever/CCC/WASH pathway although some patients with mutations in this pathway have shown dysgenesis of the corpus callosum^18,28^. These findings prompted us to investigate the cellular function of DPYSL5 in endosomal cargo retrieval and recycling.

Comparative proteomic analysis to unbiasedly quantify the interactome of wild-type DPYSL5 tagged with GFP at either its amino- or carboxy-terminal ends failed to reveal any association with proteins of the SNX17-Retriever/CCC/WASH endosomal cargo retrieval and recycling pathway (Figure S3A-S3F and Supplementary Data 2). Association of DPYSL5 with DPYSL1, DPYSL2 and DPYSL3 was consistent with their known ability to form homo-tetramers or hetero-tetramers and provided proof-of-principal validation of our interactome analysis^37–39^. A parallel analysis with the clinically identified DPYSL5-E41K and DPYSL5-G47R mutants, also failed to reveal any association with the SNX17-Retriever/CCC/WASH machinery consistent with these mutants not being a gain-of-function within this context (Figure S3A-S3F). While retaining their ability to form DPYSL5 homo- and hetero-tetramers both amino- and carboxy-terminal GFP-tagged DPYSL5-E41K, a mutation identified in eight patients with overlapping phenotypes, displayed a disrupted interaction with tropomyosin alpha-3 (TPM3), which associates with muscle and non-muscle actin. This loss in TPM3 binding was not observed with the DPYSL5-G47R mutant and hence we speculate that perturbed TPM3 interactions may be associated with DPYSL5-E41K pathogenicity.

To functionally define any role for DPYSL5 in SNX17-Retriever/CCC/WASH-mediated endosomal recycling, we generated DPYSL5 knock-out H4 neuroglioma cells. In stark contrast to the loss of cell surface ITGB1 and LRP1 observed in VPS35L, VPS26C and WASHC4 knock-out H4 cells, the loss of DPYSL5 expression had no detectable effect on surface levels of these cargo, nor did it alter the steady-state expression of Retriever, the CCC complex or the WASH complex (Figure S3G). Exogenous overexpression of wild-type GFP-DPYSL5, and GFP-DPYSL5-E41K or GFP-DPYSL5-G47R in HEK293T cells did not alter the expression of proteins in the SNX17-Retriever/CCC/WASH pathway or their cargo proteins, nor did it affect the localisation of endogenous COMMD1 to endosomes, suggesting that these mutants do not exhibit a dominant negative effect on the recycling pathway (Figure S3H and S3I). When taken together, these data suggest that mutations in DPYSL5 are likely to confer pathogenicity independent of a direct association and functional link with the SNX17-Retriever/CCC/WASH pathway. Based on these clinical and molecular data, we suggest that *DPYSL5* should not be considered a causative gene for RSS. Taken alongside the additional clinical identification of *COMMD4*, *COMMD9*, *CCDC93*, *VPS26C*, and *WASHC4* pathogenic mutations, we therefore conclude that defects in the SNX17-Retriever/CCC/WASH endosomal cargo retrieval and recycling pathway is the principal causative defect underlying RSS (Figure 2E and Figure S2F).

### Reduced cell surface expression of kidney LRP2 contributes to proteinuria in Ritscher-Schinzel syndrome

While genetic and functional evidence is consistent with RSS arising from a dysfunction in the SNX17-Retriever/CCC/WASH endosomal cargo retrieval and recycling pathway, the underlying molecular mechanism explaining why pathway dysregulation leads to the array of clinical phenotypes remains unknown. We, and others, have reported that disrupted SNX17-mediated endosomal recycling of the ΦxNPxY motif-containing LRP1 and LDLR is involved in the development of hypercholesterolemia in RSS^18,26^. Several other members of the LDL receptor-related protein (LRP) family also contain ΦxNPxY sorting motifs, and loss of these LRP family proteins have been associated with disease phenotypes (Figure S4A), of which some display phenotypic overlap with RSS^40,41^. This prompted us to investigate whether perturbed SNX17-Retriever/CCC/WASH-mediated endosomal recycling of other LRP family members may underly clinical phenotypes observed in RSS.

We first focused on LRP2, also known as megalin, as its loss of function is linked to Donnai-Barrow syndrome (MIM: 600073), a syndrome that shows neurological complications, ophthalmic anomalies such as coloboma, and low molecular weight proteinuria^42^. In the renal proximal tubules, LRP2 mediates the reabsorption of low molecular weight proteins such as β2-microglobulin, with loss of LRP2 function leading to leakage of LRP2 protein ligands into the urine^43–46^. Detailed examination of urine samples from RSS patients carrying *CCDC93* or *VPS35L* pathogenic mutations^18,19^ identified a significant increase in β2-microglobulin (Figure 3A). This prompted us to investigate whether low molecular weight proteinuria in RSS results from a loss of cell surface LRP2 expression. While previous papers suggested that LRP2 appeared not to be a SNX17 cargo^47,48^, we assessed whether LRP2 endosomal recycling is regulated by SNX17. To achieve this, we generated chimeric constructs encoding for the transmembrane domain and intracellular cytoplasmic tail of Human-LRP2 linked to a luminal facing GFP tag. As the cytoplasmic tail of LRP2 possess two ΦxNPxY sorting motifs, FENPMY^4526^ and FENPIY^4598^, we generated mutant constructs in which one or both tyrosine residues were mutated to alanine, LRP2(Y4526A), -(Y4598A), and -(Y4526A/Y4598A) (Figure 3B). Co-expression of mCherry-SNX17 with the wild-type LRP2 chimera followed by mCherry immunoisolation revealed a clear interaction between SNX17 and LRP2 that was dependent on the presence of both ΦxNPxY sorting motifs (Figure 3C). Next, we assessed the cell surface levels of endogenous LRP2 in knock-out HEK293T cell lines targeting SNX17 as well as VPS35L, CCDC22, and WASHC5^3,5^. LRP2 cell surface abundance was significantly decreased across all knock-out cell lines, which was recovered by re-expression of the corresponding wild-type proteins (Figure 3D).

**Figure 3.**
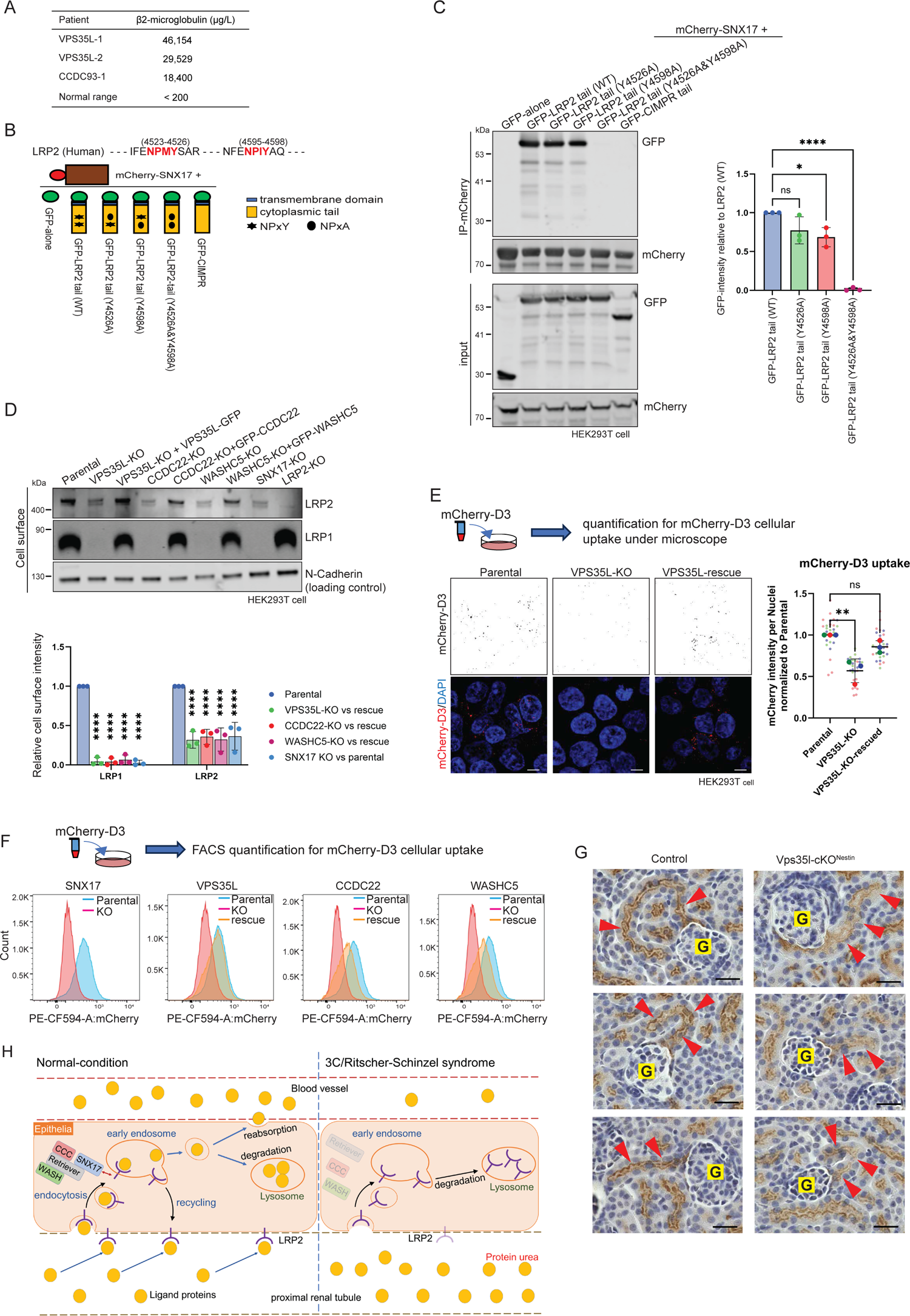
Loss of SNX17-Retriever/CCC/WASH-recycling pathway results in LRP2 dysfunction, leading to proteinuria. (A) Urinary β2-microglobulin level of individuals with biallelic pathogenic mutations in *VPS35L* or *CCDC93*. (B) The schematic illustration depicts the chimeric constructs of Human-LRP2 utilized in the study. In addition to the wild-type cytoplasmic sequence, mutant constructs were generated in which either or both NPxY motifs were substituted with NPxA. (C) Representative blots of mCherry-nanotrap for mCherry-SNX17 under co-overexpression of chimeric constructs of LRP2. GFP-tagged cytoplasmic tail of CI-MPR, a SNX-BAR cargo protein, was used as negative control. Bar graphs show band intensities relative to WT calculated from three independent experiments. (D) Representative blots for LRP1, LRP2, and N-Cadherin from three independent experiments. Cell surface protein fractions were obtained from HEK293T cell lines. Bar graphs show relative values of KO cells to their rescue or parental cells. (E) Representative view of mCherry-D3 uptake in parental, VPS35L-KO, and VPS35L-rescue cells. Cells were incubated with mCherry-D3 for 30 min, followed by DAPI staining and imaging with a fluorescence microscope. mCherry intensity was quantified using Image J software. 10 fields were acquired in each condition in each of three independent experiments, and mCherry intensity of each field was normalized to the number of DAPI-stained nuclei. Scale bars, 10 µm. (F) HEK293T cell lines were incubated with mCherry-D3 for 30 min followed by FACS analysis to quantitate cellular uptake of mCherry-D3. (G) Immunohistochemistry of Lrp2 in renal tissue in littermate control or Vps35l-cKO^Nestin^. Red arrows indicate Lrp2 in S1 segment of proximal tubules. Three mice were analyzed in each group. G; Glomerulus. (H) Schematic illustration of the molecular mechanism underlying the proteinuria observed in Ritscher-Schinzel syndrome. (C, D, E) Error bars represent mean± SD. *, P<0.05; **, P<0.01; ***, P<0.001; ****, P<0.0001.

To functionally define the role of LRP2 in the uptake of extracellular protein ligands, we bacterially expressed and then purified an mCherry-tagged D3 domain of Receptor Associated Protein (RAP-D3), a well-established LRP2 ligand^49^. Using confocal microscope and FACS analysis in the knock-out cells, we observed a significant reduction in the cellular uptake of RAP-D3 that was rescued by re-expression of the corresponding proteins (Figure 3E and 3F). Finally, to confirm the functional requirement of the SNX17-Retriever/CCC/WASH pathway in LRP2 endosomal recycling *in vivo* we generated a flox-*Vps35l* mouse model since a germline knock-out of Vps35l induced embryonic lethality during mid-gestation^19^. We crossed this line with Nestin-Cre mice to knock-out Vps35l expression in proximal tubules of the kidney and a range of central nervous system tissues (Figure S4B). Loss of Vps35l expression and reduced Lrp1 expression was confirmed by western analysis of mouse brain homogenates (Figure S4C). While PAS staining and ultrastructural analysis revealed a normal renal structure in Vps35l-cKO^Nestin^ mice, immunohistochemistry of renal sections showed a reduced LRP2 expression especially in the S1 segment of proximal tubules when compared to their litter mate controls (Figure 3G, S4D-F). These *in vivo* data confirm that the SNX17-Retriever/CCC/WASH pathway is required for the proper recycling of LRP2 in the kidney. Together these findings establish that the cell surface level of LRP2 is regulated by the Retriever/CCC/WASH cargo recycling pathway through SNX17 recognition of its ΦxNPxY sorting motif. In RSS patients, we propose that disruption of LRP2 recycling results in reduced cell surface expression of LRP2 leading to reduced protein ligand uptake in the proximal renal tubule and the development of the clinically observed proteinuria (Figure 3H).

### SNX17-Retriever/CCC/WASH pathway orchestrates retrieval and recycling of membrane proteins important for skeletal development

Skeletal malformation is one of the core phenotypes observed in RSS. Another member of the lipoprotein receptor family, LRP4 is known to play an essential role in the regulation of Wnt signalling during development of the skeletal system and it possesses a ΦxNPxY motif in its cytoplasmic tail (YSNPSY^17^^69^) - mutations in LRP4 are causative for congenital malformation Cenani-Lenz syndactyly syndrome, which affects bone development in the hands, forearms and lower limbs^50–53^. To probe its potential role as a cargo for SNX17-mediated endosomal recycling, we again engineered chimeric constructs encoding for the transmembrane domain and intracellular cytoplasmic domain of Human-LRP4 linked to a luminal facing GFP tag. In co-expression studies alongside mCherry-SNX17 in HEK293T cells, mCherry-trap immuno-isolation revealed wild-type LRP4 binding to SNX17 that was lost in a corresponding LRP4(Y1769A) mutant. Moreover, in a series of human H4 neuroglioma cell lines knocked-out for the expression of SNX17, VPS35L, CCDC22, or WASHC5, the cell surface level of LRP4 was significantly reduced and was rescued upon re-expression of the corresponding wild-type proteins (Figure 4A and 4B).

**Figure 4.**
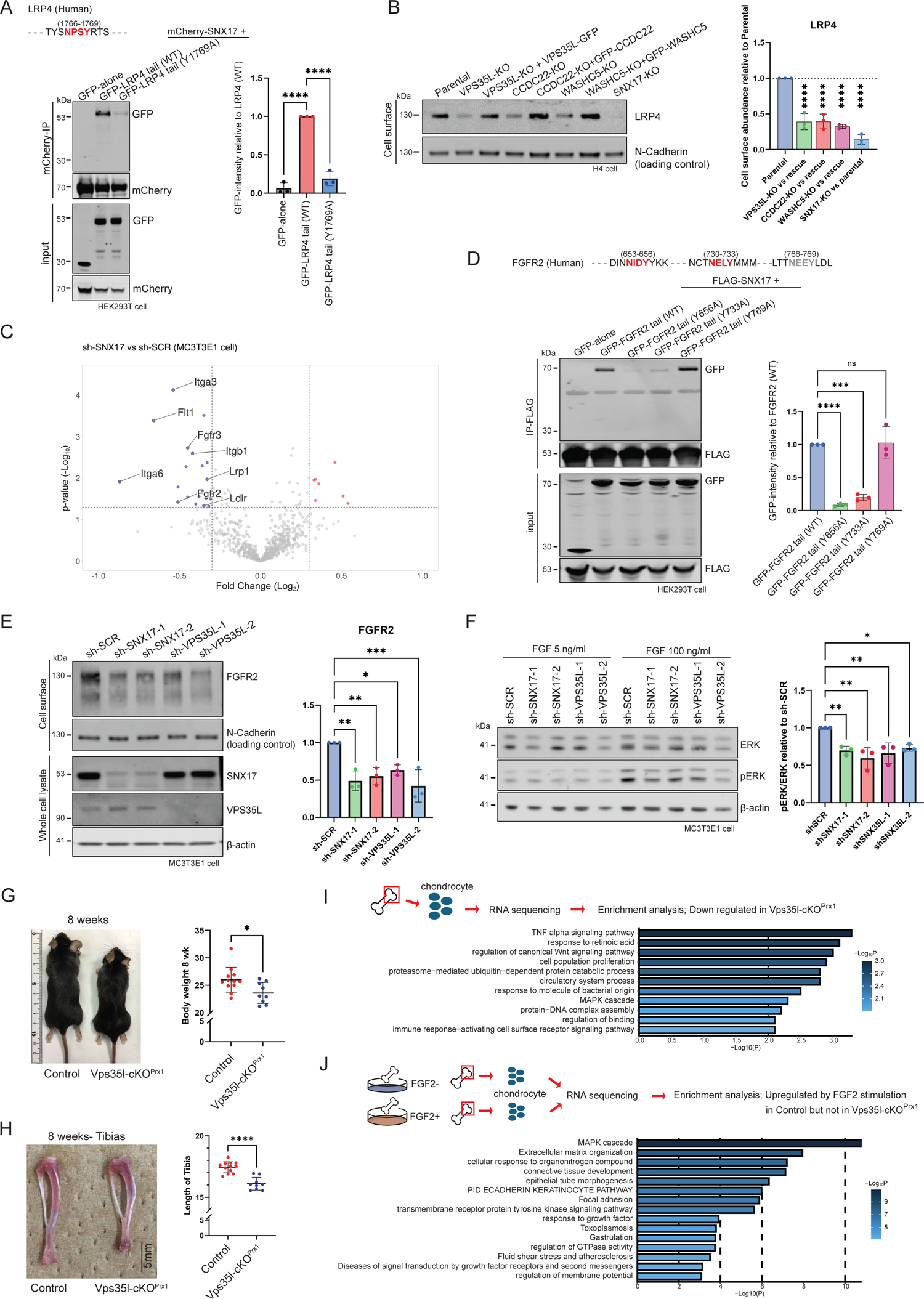
Loss of membrane proteins such as LRP4 and FGFR2 involved in disease pathogenesis in skeletal system in Ritscher-Schinzel syndrome. (A) Representative blots of Co-expression of mCherry-tagged SNX17 with GFP-tagged cytoplasmic tail of Human-LRP4 from three independent experiments. The interaction between mCherry-SNX17 and wild-type GFP-tagged cytoplasmic tails of LRP4 was significantly decreased when NPxY was substituted with NPxA. Bar graphs show relative density of the bands in immunoprecipitated proteins from three independent experiments. (B) Representative blots of LRP4 in cell surface protein fraction obtained from H4 cell line. N-Cadherin was used for loading control. Bar graph shows relative intensity of knock-out cells to their rescue or parental cells among three independent experiments. (C) Volcano plots of transmembrane proteins with decreased (blue circles) or increased (red circles) cell surface abundance in TMT-based proteomics in sh-SNX17 comparing to sh-SCR in MC3T3-E1 mouse osteoblast cells from three independent experiments. Two independent sh-RNAs for SNX17 were used to avoid off-target effects. (D) Representative blots for FLAG-nanotrap of FLAG-SNX17 under co-overexpression of chimeric constructs of FGFR2 cytoplasmic tail from three independent experiments. NxxY motifs were mutated to NxxA in the mutant. (E) Representative blots for FGFR2 in MC3T3-E1. Cell surface protein fraction was obtained, and N-Cadherin were used for loading control. Bar graphs show relative values of sh-SNX17 or sh-VPS35L knock-down cells compared to sh-SCR control cell (n=3). (F) Representative blots for ERK and pERK under stimulation of FGF2 in MC3T3-E1 from three independent experiments. Cells were cultured with 5ng/ml of FGF-2 overnight, then media was replaced with fresh media with 100ng/ml of FGF-2 for 7 min. Cells were lysed and analysed by western blotting. (G) Representative pictures and scatter plots of their body weight in VPS35L^Prx1^-cKO mice and their littermate controls. [Control; n=12, Vps35l-cKO^Prx1^; n=9]. (H) Representative images and graph showing length of tibiae in mice at 8 weeks with indicated genotype. [Control; n=12, Vps35l-cKO^Prx1^; n=9]. (I) Gene enrichment analysis of downregulated genes in Vps35l-cKO^Prx1^ compared to their littermate controls was performed using RNA sequencing analysis data. Total RNA was extracted from E16.5 mice chondrocytes. [Control; n=5, Vps35l-cKO^Prx1^; n=3]. (J) Gene enrichment analysis of gene sets, where genes upregulated by FGF2 stimulation in littermate controls but not in Vps35l-cKO^Prx1^ were included. Tibiae obtained from E16.5 mouse embryos of either Vps35l-cKO^Prx1^ or their littermate controls were cultured for four days with or without FGF2, followed by RNA extraction for RNA sequencing analysis. [Control; n=3, Vps35l-cKO^Prx1^; n=4]. (A, B, D, E, F) Error bars represent mean ± SD. *, P<0.05; **, P<0.01; ***, P<0.001; ****, P<0.0001.

We then contemplated whether other proteins, beyond the LRP family, could be regulated by the SNX17-Retriever/CCC/WASH recycling axis, and whether their loss of recycling contributes to skeletal complications in RSS. To this end, we used cell surface proteome analysis as an unbiased and quantitative methodology. Two independent shRNA were employed to suppress SNX17 in MC3T3E1 cells, a mouse osteoblast cell line, and then cell surface proteins were biotinylated to isolate a cell surface fraction through streptavidin affinity capture. A total of 884 transmembrane proteins were identified of which 16 were lost significantly by more than 20% from the SNX17 suppression when compared with controls (Figure 4C and Supplementary Data 3). Out of those proteins, LRP1, ITGB1, and LDLR all contain ΦxNPxY motifs in their cytoplasmic tails, whilst FGFR2 and FGFR3 contain ΦxNxxY motifs. FGFR2 and FGFR3 belong to FGF receptor family, which play pivotal roles in skeletal development, and are genetically implicated in human skeletal disorders^54^. We therefore, investigated the association between SNX17 and FGFR2, a receptor whose cytoplasmic domain contains one ΦxNxxY and two NxxY motifs (INNIDY^656^, and CTNELY^733^ and TTNEEY^769^ respectively). Immunoprecipitation and quantitative western analysis of the GFP-tag transmembrane and cytoplasmic facing tail of Human-FGFR2 co-expressed with FLAG-tagged SNX17 identified the association of SNX17 with wild-type FGFR2, an interaction that was significantly reduced in FGFR2-(Y656A) and -(Y733A) mutants (Figure 4D). Consistent with a functional role for SNX17 association with FGFR2, the cell surface abundance of this receptor was decreased in both SNX17- and VPS35L-suppressed MC3T3E1 cells when compared to control cells, and this reduction resulted in attenuated FGF-mediated signaling through the MAPK pathway (Figure 4E and 4F).

To further confirm the contribution of SNX17-Retriever/CCC/WASH function in skeletal development, we utilized our mouse model of RSS. Flox-Vps35l mice were crossed with Prx1-Cre mice to ablate Vps35l expression in the skeletal system. Consistent with RSS patients displaying growth impairment and short stature, Vps35l-cKO^Prx1^ mice showed a smaller body weight and shorter length of tibias (Figure 4G and 4H). To elucidate the molecular mechanism underlying this RSS-like skeletal phenotypes, we extracted mRNA from chondrocytes in E16.5 mouse tibiae and performed RNA sequencing analysis. Gene enrichment analysis of downregulated genes in Vps35l-cKO^Prx1^ compared to litter mate controls (Fold change < −0.585, q < 0.1) revealed enrichment of gene sets in the gene ontology (GO) term “regulation of canonical Wnt signaling pathway” and “MAPK cascade” suggesting these pathways were dysregulated potentially due to loss of LRP4 and FGFR2, respectively (Figure 4I). These observations prompted further investigation into whether FGF signalling is disrupted in our *in vivo* model. In an *ex vivo* tibiae culture system, we cultured tibiae obtained from E16.5 mice in medium with or without FGF2 for four days, and then extracted RNA for sequence analysis. Gene enrichment analysis of upregulated genes (Fold change > 0.585, q < 0.05) by FGF2 stimulation in littermate controls, but not in Vps35l-cKO^Prx1^, revealed that gene sets related to the “MAPK cascade” were the top hit, consistent with significant disruption of FGF signalling in developing bone of Vps35l-cKO^Prx1^ mice (Figure 4J and S5A-F). Overall, these data indicate that the SNX17-Retriever/CCC/WASH cargo retrieval and recycling pathway is essential for skeletal development through maintenance of cell surface levels of proteins such as LRP4 and FGFR2, that are essential for normal bone development through Wnt and MAPK signaling cascades.

### SNX17-Retriever/CCC/WASH dependent retrieval and recycling regulates cargos essential for neuronal development

Next, we investigated the role of SNX17-Retriever/CCC/WASH pathway in neuronal development, another hallmark of RSS. We first focused on interactions between SNX17 and two LRP family proteins, LRP8 and VLDLR. Both contain ΦxNPxY motifs in their cytoplasmic tails, FDNPVY^866^ and FDNPVY^837^ respectively, and they are each essential for neurodevelopment as receptors for Reelin signalling^55–57^. Expression of the GFP-tagged chimeric protein encoding the transmembrane and cytoplasmic tails of wild-type Human-LRP8 and Human-VLDLR showed clear interactions with mCherry-SNX17, which were significantly reduced in corresponding LRP8(Y866A) and VLDLR(Y837A) mutants (Figure 5A). Consistent with the endosomal recycling of LRP8 and VLDLR being regulated by the SNX17-Retriever/CCC/WASH pathway, cell surface abundance of each receptor was significantly reduced in primary neurons under suppression of SNX17 or the Retriever subunit VPS35L (Figure 5B). Suppression of SNX17 or VPS35L in primary rat cortical neurons also resulted in a decrease in the uptake of Reelin (Figure 5C), and consequently a reduction in Reelin-mediated phosphorylation of Dab1, a downstream phosphorylation target of Reelin signalling (Figure 5D).

**Figure 5.**
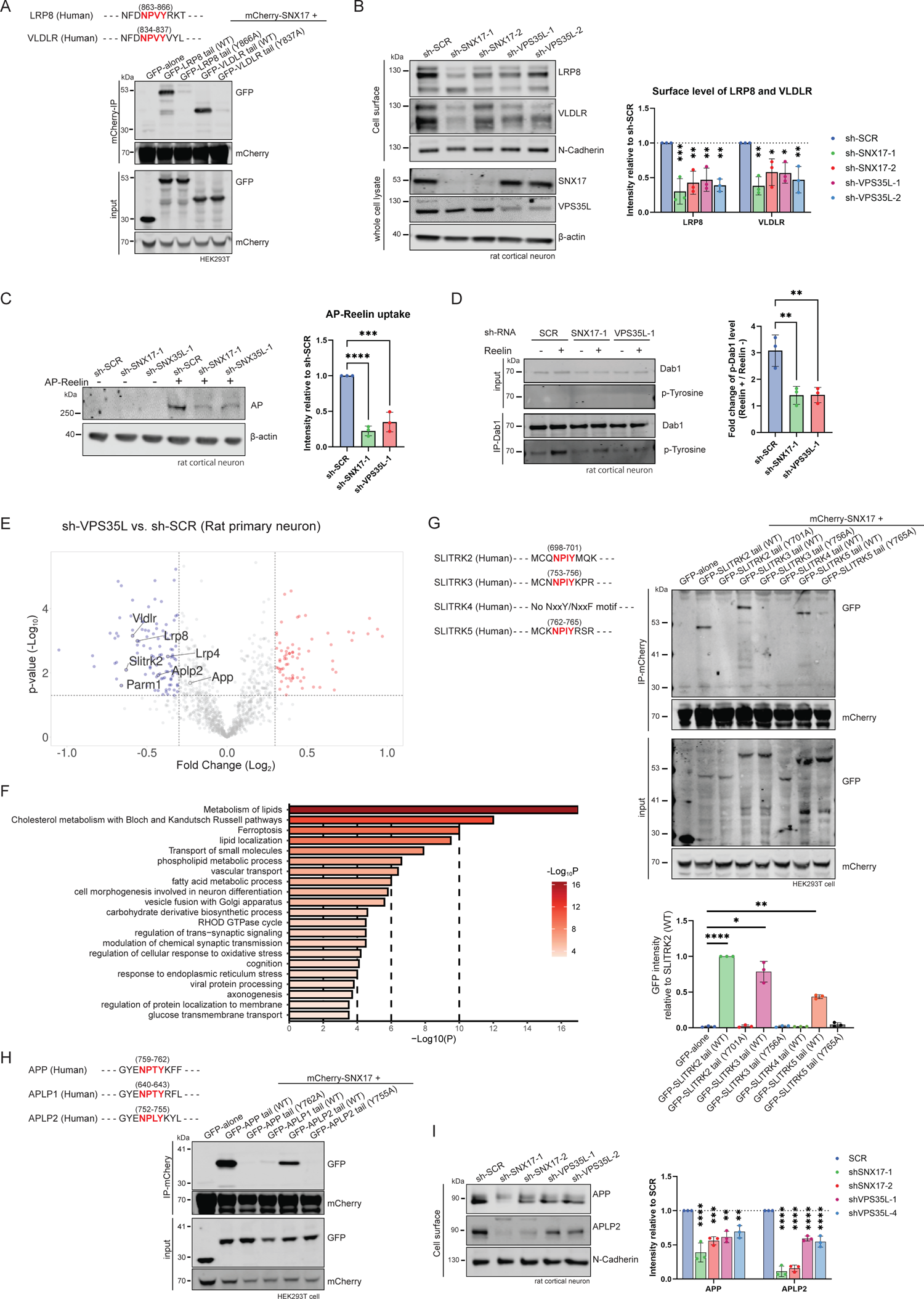
SNX17-Retriever/CCC/WASH pathway is essential for recycling of Reelin signaling receptors, APP family, and SLITRK family proteins. (A) Representative blots of co-expression of mCherry-tagged SNX17 with GFP-tagged cytoplasmic tail of LRP8 and VLDLR from three independent experiments. The interactions between mCherry-SNX17 and wild-type GFP-tagged cytoplasmic tails of Human-LRP8 and Human-VLDLR were significantly decreased when NPxY was substituted with NPxA. (B) Representative blots of analysis from three independent experiments using DIV17 rat cortical neurons transduced with either a scramble-control, SNX17, or VPS35L shRNA. Bar graphs show relative protein abundance of LRP8 and VLDLR in cell lysate or cell surface. (C) DIV17 rat cortical neurons transduced with shRNA were incubated for 30 min with or without AP-Reelin, followed by cell lysis and western blot analysis. Bar graph shows quantification of band intensities of AP relative to cells transduced with sh-SCR control from three independent experiments. (D) DIV17 rat cortical neurons transduced with shRNA were incubated for 7 min with or without Reelin. Phosphorylation level of Dab1 was then measured using immunoprecipitation and western blot analysis. Representative blots and quantification from three independent analyses are shown. (E) Volcano plots of transmembrane proteins with decreased (blue circles) or increased (red circles) cell surface abundance in sh-VPS35L suppression comparedto sh-SCR in DIV17 rat cortical neuron from three independent TMT-based proteomic experiments. Two independent sh-RNAs for VPS35L were used to avoid off-target effects. (F) Enrichment analysis of significantly downregulated proteins (LogFC < −0.32, p < 0.05) in the sh-VPS35L compared to sh-SCR by Metascape. (G) Representative blots of mCherry-nanotrap for mCherry-SNX17 under co-overexpression of chimeric constructs of Human-SLITRK family proteins from three independent experiments. All proteins except for SLITRK4 have an NPxY motif, which was mutated to NPxA in the mutant. (H) Representative blots of mCherry-nanotrap for mCherry-SNX17 under co-overexpression of chimeric constructs of Human-APP family proteins from three independent experiments. APP, APLP1, and APLP2 have NPxY motifs, and the NPxY motif was mutated to NPxA in the mutant. (I) Representative blots for APP and APLP2 in rat cortical neurons. Cell surface protein fraction was obtained, and N-Cadherin were used for loading control. Bar graphs show relative values of cells with sh-SNX17 or sh-VPS35L suppression compared to sh-SCR control cells (n=3). In all graphs, error bars represent mean ± SD. *, P<0.05; **, P<0.01; ***, P<0.001; ****, P<0.0001.

To identify additional integral membrane proteins that potentially may contribute to the complex neurological phenotypes observed in RSS upon perturbation of the SNX17-Retriever/CCC//WASH pathway, we again utilized unbiased and quantitative cell surface proteomics. For this analysis, VPS35L was suppressed in rat cortical primary neurons through the transduction of two independent shRNAs, and then cell surface proteins were biotinylated to isolate a cell surface fraction through streptavidin affinity capture. Among over 5,500 identified proteins in TMT-based quantitative proteome analysis, the cell surface expression of 99 transmembrane proteins were significantly reduced by more than 20% in VPS35L suppressed cells when compared to controls (Figure 5E and Supplementary Data 4). Gene ontology enrichment analysis of downregulated genes (LogFC < −0.32 and p < 0.05) revealed terms including “metabolism of lipids”, “Cholesterol metabolism with Bloch and Kandutsch Russel pathways”, and “lipid localisation”, suggestive of neuronal metabolic alterations, potentially reflecting dysregulation of LRP family proteins (Figure 5F). Out of those proteins significantly decreased in the cell surface proteome following VPS35L-suppression, APLP2, LRP8, VLDLR, LRP4, PARM1, and SLITRK2 all contain NPxY sorting motifs in their cytoplasmic domains.

SLITRK2 is genetically linked to neurodevelopmental disorders and is one member of the SLITRK protein family that have been implicated in synapse development^58,59^. Chimeric proteins encoding for the cytoplasmic tails of Human-SLITRK2, Human-SLITRK3, and Human-SLITRK5, each of which contains an NPxY motif, CQNPIY^701^, CNNPIY^756^, and CKNPIY^765^ respectively, revealed an interaction with mCherry-SNX17 that was lost with the corresponding NPIA mutants – as a negative control we utilized the cytoplasmic tail of Human-SLITRK4 that lacks an NPxY motif (Figure 5G). These NPxY-containing SLITRK proteins therefore have the hallmarks of cargos for neuronal SNX17-Retriever/CCC/WASH pathway retrieval and recycling.

Another identified protein potentially involved in RSS neuronal disease pathology is APLP2, a member of the APP protein family along with APP and APLP1. While APP is a well-known protein associated with neurodegenerative disorders, the other APP family proteins are also believed to be crucial for neurodevelopment, for example depletion of APP and APLP2 leads to embryonic lethality in mice^60–63^. All three proteins contain the ΦxNPxY motif in their cytoplasmic domains (APP - YENPTY^762^, APLP1 - YENPTY^643^, APLP2 - YENPTY^755^) and previous reports have indicated that APP is a cargo reliant on SNX17 for its endosomal retrieval and recycling^64^. This prompted us to investigate whether APP proteins can be retrieved and recycled by the SNX17-Retriever/CCC/WASH pathway. Chimeric proteins expressing the GFP-transmembrane-cytoplasmic domain of Human-APP or Human-APLP2 showed an ΦxNPxY-dependent interaction with mCherry-SNX17 – under these conditions no association was observed with Human-APLP1 (Figure 5H). Analysis of the cell surface levels of APP and APLP2 using surface restricted biotinylation and quantitative western blot analysis in cultured primary rat cortical neurons, confirmed that the levels of APP and APLP2 were both significantly reduced under SNX17 and VPS35L suppression (Figure 5I). Together, these unbiased data revealed that alterations in the cell surface expression of several integral membrane proteins, that include Reelin signalling receptors, SLITRK proteins and APP family members, are likely to contribute to the neurodevelopmental pathology and neurological complications observed in RSS.

To further probe the complexities of RSS associated neuropathology, we turned to an *in vivo* analysis using our Vps35l-cKO^Nestin^ mice, since Nestin expression is observed in many neuronal cell types. Almost 30% (23/72) of Vps35l-cKO^Nestin^ mice showed profound hydrocephalus, an observed neurological complication in RSS (Figure 6A-6C), although thickness of brain cortex, and the migration and number of neurons appeared not to be significantly affected in the brains of mice lacking profound hydrocephaly (Figure 6D and S6A-S6B). Regardless of the presence or absence of hydrocephalus, all mice showed growth impairment, behavioural problems, and high mortality rates (Figure 6E, 6F, and Supplementary Movies). These phenotypes are consistent with our Vps35l-cKO^Nestin^ mouse model recapitulating many of the neurological phenotypes observed in RSS patients. As our cell surface proteomic analysis in rat cortical neurons suggested that proteins involved in the “modulation of chemical synaptic transmission” are affected under VPS35L suppression (Figure. 5F), we investigated the alterations of membrane proteins and synaptic proteins at the synapse using this Vps35l-cKO^Nestin^ mice model.

**Figure 6.**
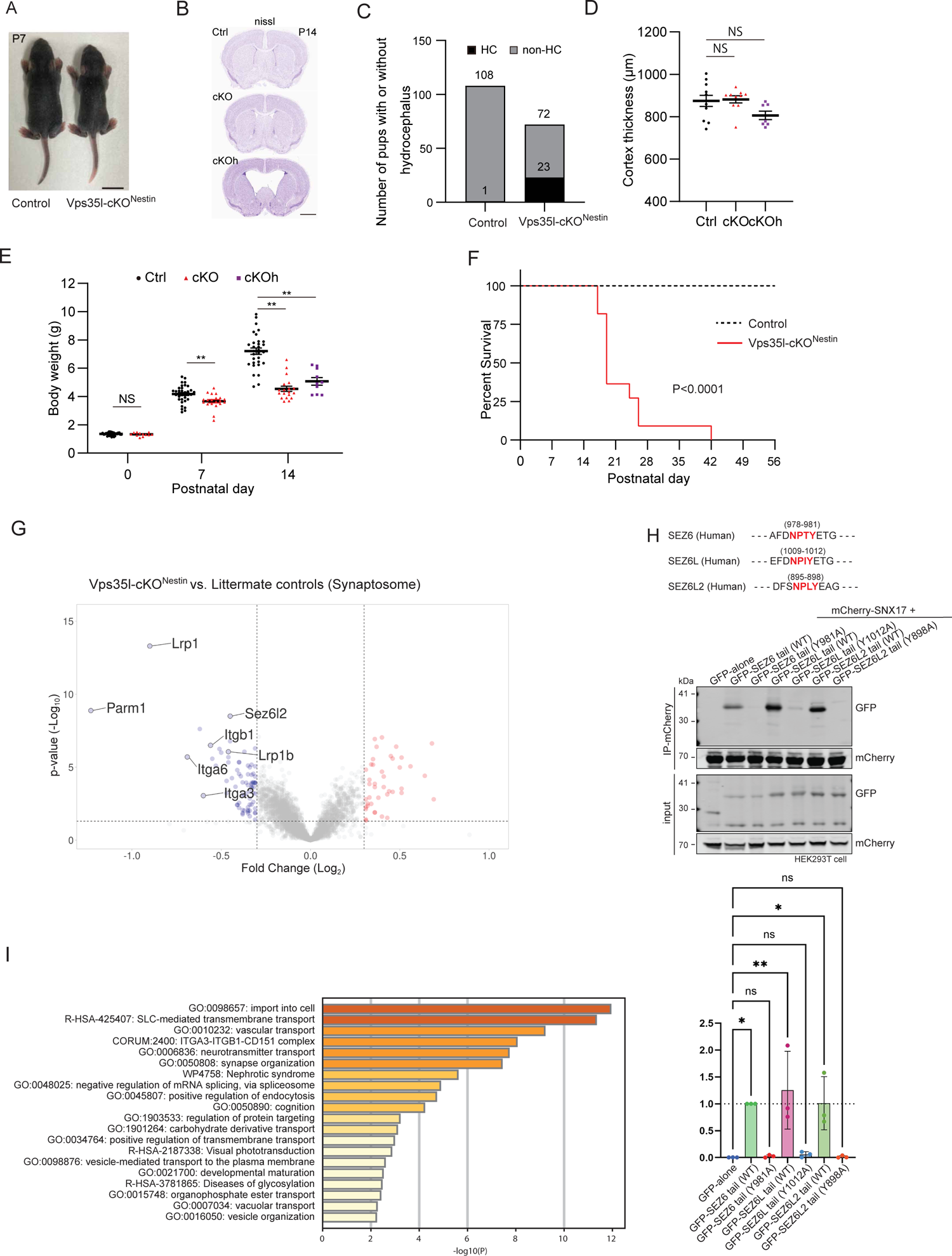
Loss of SNX17-Retriever/CCC/WASH recycling pathway causes severe synaptic effects. (A) Representative pictures of Vps35l-cKO^Nestin^ and their littermate controls at postnatal day 7 (P7). Scale bar, 1 cm. (B) Nissl staining of P14 brain slices from mice with indicated genotypes. VPS35L-cKO^Nestin^ mice were divided into two groups: those without gross hydrocephalus (cKO), and those with gross hydrocephalus (cKOh). Slices at anterior level are shown. Scale bar, 1 mm. (C) Bar graph depicting the number of mice with or without gross hydrocephalus. “HC” indicates gross hydrocephalus, while “non-HC” represents those without obvious hydrocephalus. (D) Scatter plots of cortex thickness of Vps35l-cKO^Nestin^ mice and their littermate controls at P14. [Ctrl; n=11, cKO; n=10, cKOh; n=7]. (E) Scatter plots of their body weight are shown. [P0; Ctrl; n=32, cKO; n=12, P7; Ctrl; n=27, cKO=23, P14; Ctrl; n=53, cKO; n=19, cKOh: n=10]. (F) Caplan-Meier curves with log-rank (Mantel-Cox) test show a significant decrease in the survival ratio of Vps35l-cKO^Nestin^ compared with control mice [n=11 in each group]. (G) Volcano plots of transmembrane and synaptic proteins with decreased (blue circles) or increased (red circles) abundance in Vps35l-cKO^Nestin^ mice compared to their litter mate controls. Synaptic protein fractions were isolated from hippocampal neurons of P7 mice, followed by TMT-based proteome analysis. [Control; n=8, Vps35l-cKO^Nestin^; n=6]. (H) Representative blots of mCherry-nanotrap for mCherry-SNX17 under co-overexpression of chimeric constructs of Human-SEZ6 family proteins from three independent experiments. All proteins have an NPxY motif, which was mutated to NPxA in the mutant. (I) Enrichment analysis by Metascape. Proteins significantly affected in Vps35l-cKO^Nestin^ (|LogFC| > 0.32, p < 0.05) with GO term with “Synapse” and/or “Transmembrane” were included into the analysis. In all graphs, error bars represent mean ± SD. *, P<0.05; **, P<0.01.

Synaptic fractions were obtained from the mouse hippocampus at P7 using a standard synaptosome isolation method (Figure S6C)^65^. The enrichment of synaptic proteins was confirmed by the significant accumulation of PSD95 in the synaptosome fraction when compared to the total lysate (Figure S6D). Proteomic analysis of this synaptosome fraction confirmed the alteration of membrane proteins in the mouse synapse. Consistent with those membrane proteins identified as requiring the SNX17-Retriever/CCC/WASH pathway in our cultured cell model, many proteins were found to be decreased in Vps35l-cKO^Nestin^ mouse hippocampal synapses, including lipoprotein receptors, integrins and PARM1 (Figure 6E and Supplementary Data 5). In addition, SEZ6L2 (anti-seizure-related 6 homolog like 2), one of the SEZ6 family of proteins reported to be essential for fetus development and linked to cerebellar ataxia^66–68^, was also decreased in our synaptosome fraction. As the cytoplasmic domains of SEZ family proteins, Human-SEZ6, Human-SEZ6L and Human-SEZ6L2 contain ΦxNPxY motifs (FDNPTY^981^, FDNPIY^1012^, FSNPLY^898^ respectively), we generated GFP-tagged wild-type and mutant constructs where each ΦxNPxY was substituted to ΦxNPxA. When co-overexpressed alongside mCherry-SNX17, all wild-type cytoplasmic tail domains showed binding to SNX17 that was lost with the corresponding ΦxNPxA mutant (Figure 6H). Cell surface restricted analysis confirmed that SEZ6L2 levels were reduced in all VPS35L, CCDC22, and WASHC5 knock-out HEK293T cells lines (Figure S7A, S7B and Supplementary Data 6). Enrichment analysis from our analysis of Vps35l-cKO^Nestin^ mouse hippocampal synapses, also indicated the dysregulation in gene sets with GO terms such as “neurotransmitter transport” and “synapse organization” (Figure 6I). Additionally, gene sets with GO terms such as “visual phototransduction” and “cognition” were dysregulated, potentially reflecting the clinical phenotype of cognitive disorders and ophthalmological complications documented in RSS patients. In summary, these data establish that the SNX17-Retriever/CCC/WASH recycling pathway regulates a variety of cargo proteins in the neuronal system, including integrins, SLITRK and SEZ6 protein families, Reelin receptors and APP proteins, and that dysregulation in the cell surface expression of these proteins likely explains the highly complex neurodevelopmental pathology observed in RSS.

## Discussion

In this study we have combined clinical and genetic analyses with cellular and biochemical approaches, including unbiased proteomics and *in vivo* analysis in mice, to define that dysregulation of cargo retrieval and recycling through the SNX17-Retriever/CCC/WASH pathway is causative for RSS and to identify a spectrum of integral membrane proteins whose cell surface expression is reduced in key tissues affected in RSS. In so doing, we have revealed the mechanistic basis for the correct development and function of the kidney, bone and brain that likely underly the clinical manifestations observed in patients with RSS. To the best of our knowledge, this is the first report combining unbiased proteome analysis of multiple tissue-derived cell lines with pathological analysis in a mouse disease model to understand the fundamental molecular mechanisms underlying a congenital human malformation syndrome that affects multiple organs. Collectively, our work defines that perturbed sorting of tissue-specific cargo proteins through the SNX17-Retriever/CCC/WASH endosomal recycling pathway is the principal root cause of RSS. Perturbed recycling reduces the cell surface expression of specific cargo proteins essential for signalling and cell-cell communications necessary for the development of multiple human tissues. We therefore consider that RSS constitutes an endosomal “recyclinopathy”.

Cellular and mouse histopathological analyses has revealed that impairment of endosomal recycling of LRP2, and the resulting reduction of its cell surface abundance, contributes to the development of proteinuria observed in RSS, wherein low molecular weight LRP2 ligands such as β2-microglobulin leak into the urine. Perturbed recycling and ensuing reduced cell surface expression of other LRP family proteins, including LRP4, LRP8, and VLDLR, are likely to contribute to the development of skeletal and neurological complications, given that their functions are essential for the development of these tissues^50,51,55,69^. Moreover, unbiased approaches using several different cell lines also identified proteins, including FGFR2, and members of the APP, SLITRK, and SEZ6 protein families, as cargo proteins for recognition by SNX17 and sorting through the Retriever/CCC/WASH retrieval and recycling pathway. Considering the already reported importance of these proteins in human development and their dysregulation in human disease (Figure S8)^42,51,52,55,58–61,68,70^, our findings strongly suggest that their reduced SNX17-Retriever/CCC/WASH-mediated recycling through the endosomal network and accompanying reduction in cell surface expression further underlies the phenotypes observed in the skeletal and nervous systems of RSS patients. While further investigation is needed to determine the specific impact of dysfunction in each protein on disease development, the establishment of the concept of membrane protein retrieval and recycling abnormalities based on our experimental data paves the way for consideration of diagnostic and potential therapeutic interventions to aid those symptoms experienced by RSS patients.

SNX17 has been suggested to be crucial for synaptic function through the recycling of ITGβ1^71^. Our unbiased analysis using cell lines and a VPS35L-cKO mouse model is consistent with the molecular pathophysiology of each tissue, particularly the nervous system, being complex and mediated through perturbed endosomal sorting of numerous cargo proteins. The phenotypes observed in RSS are therefore the result of accumulated changes in the expression of functionally diverse integral membrane proteins rather than a single protein.

While we have identified several novel genes responsible for RSS, further accumulation of genetic evidence is still required. The identification of further patients will provide a greater understanding of the clinical diversity of RSS and the specific impact of each protein in the SNX17-Retriever/CCC/WASH pathway. Although the COMMD9 variant identified in this study resulted in a premature stop codon, COMMD9 knock-out cells displayed a relatively milder reduction in cargo protein abundance compared to knock-out cells of COMMD4 or CCDC93, whose identified mutations retained the ability to form the CCC complex. These findings suggest that residual function within the SNX17-Retriever/CCC/WASH pathway may influence the diversity of complications and their severity among patients with RSS. The role of different genetic backgrounds also needs to be considered when questioning pathogenic severity. In this regard, patients harbouring the COMMD4-L41R mutation were born to consanguineous parents and all three patients passed away during early childhood (ages 0-5). This suggests that this severe form of RSS may arise from an additional compounding disease-causing gene mutation(s) that has yet to be identified.

In sum, we have defined that the multiple organ defects observed in RSS arise from a common perturbation in the assembly and stability of the Commander super-assembly that leads to a common defect in the endosomal retrieval and cell surface recycling of key cargo proteins that perform essential functions in the development of those organs and tissues clinically affected in RSS. As such we propose that RSS constitutes an endosomal recyclinopathy.

### Limitations of the study

While our work has identified additional mutations in the Commander pathway causative for RSS the genetic identification and clinical characterisation of additional RSS patients will be essential to establish a fully detailed genotype:phenotype relationship. In addition, although we have defined how perturbed SNX17-Retriever/CCC/WASH endosomal recycling leads to tissue-specific alterations in the cell surface proteome that likely underlies the defects observed in kidney, bone, and brain development, further studies will be required to establish the defects associated with other RSS phenotypes including cardiac defects and cerebellar dysplasia. Our functional analysis of the SNX17-Retriever/CCC/WASH pathway has been restricted to effects at the cell surface. An equivalent analysis utilising proteomics restricted to other organelles, for example the Golgi apparatus and lysosomes, may be required to generate a greater holistic view of defective endosomal retrieval and recycling in RSS. Finally, we have primarily focused on analysing cargo proteins regulated by SNX17, but the unbiased proteome analysis also identified expression changes in various membrane proteins that do not contain NxxY/F motifs. Understanding how these membrane proteins are directly or indirectly regulated by the Retriever/CCC/WASH pathway will further our comprehension of the overall molecular pathology of RSS.

## Data Availability

All data produced in the present study are available upon reasonable request to the authors.

---

**Figure S1.**
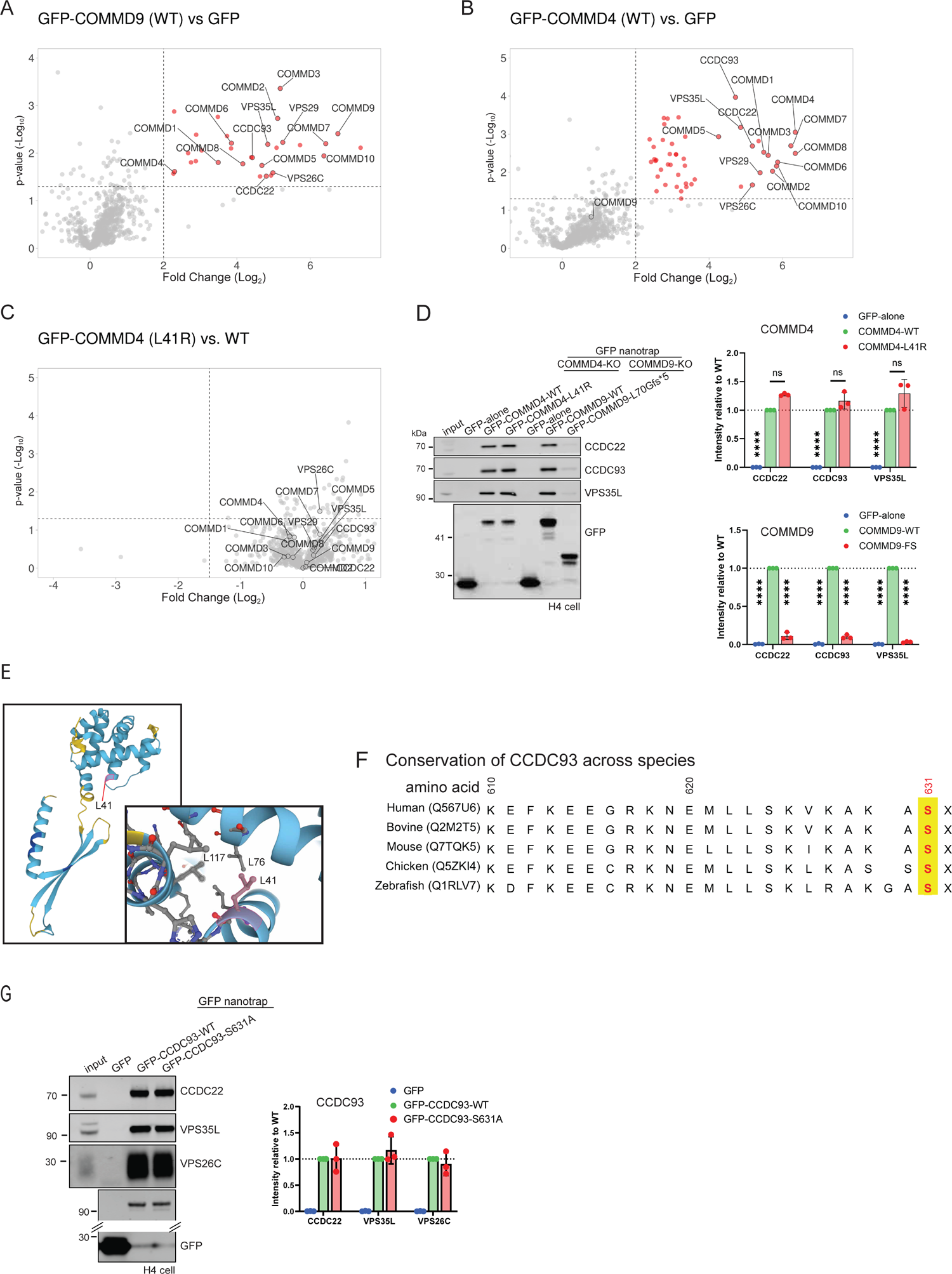
Functional analysis of identified mutations in COMMD4, *COMMD9*, and CCDC93 (A) Volcano plots of enriched (red circles) interactors in GFP-COMMD9-WT (wild-type) compared to GFP-alone in TMT-based proteomics with three replicates. (B and C) Volcano plots of enriched (red circles) and decreased (blue circles) interactors in TMT-based proteomics with three replicates. The comparisons include: GFP-COMMD4-WT vs. GFP-alone, and GFP-COMMD4-L41R vs. GFP-COMMD4-WT. (D) Representative blots of GFP-nanotrap analysis for COMMD4 and COMMD9. Wild-type or identified variants of GFP-COMMD4 or GFP-COMMD9 fusion protein was expressed in their knock-out H4 cell line. Bar graphs show relative density of the bands in immunoprecipitated proteins from three independent experiments. (E) AlphaFold2 model of COMMD4 with Leu41 indicated in the model. Close-up view of the mutation site suggests Leu41 forms hydrophobic pocket with highly conserved residues of L76 and L117. (F) Sequence alignment of CCDC93 across species. The mutated amino acid residue of Ser631 is marked by red and yellow. (G) Representative blots of GFP-nanotrap analysis for CCDC93. Wild-type or S631A variant of GFP-CCDC93 fusion protein was expressed in H4 CCDC93-KO cell line. Bar graphs show relative density of the bands in immunoprecipitated proteins from three independent experiments. (D and G) Error bars represent mean± SD. *, P<0.05; **, P<0.01; ***, P<0.001; ****, P<0.0001.

**Figure S2.**
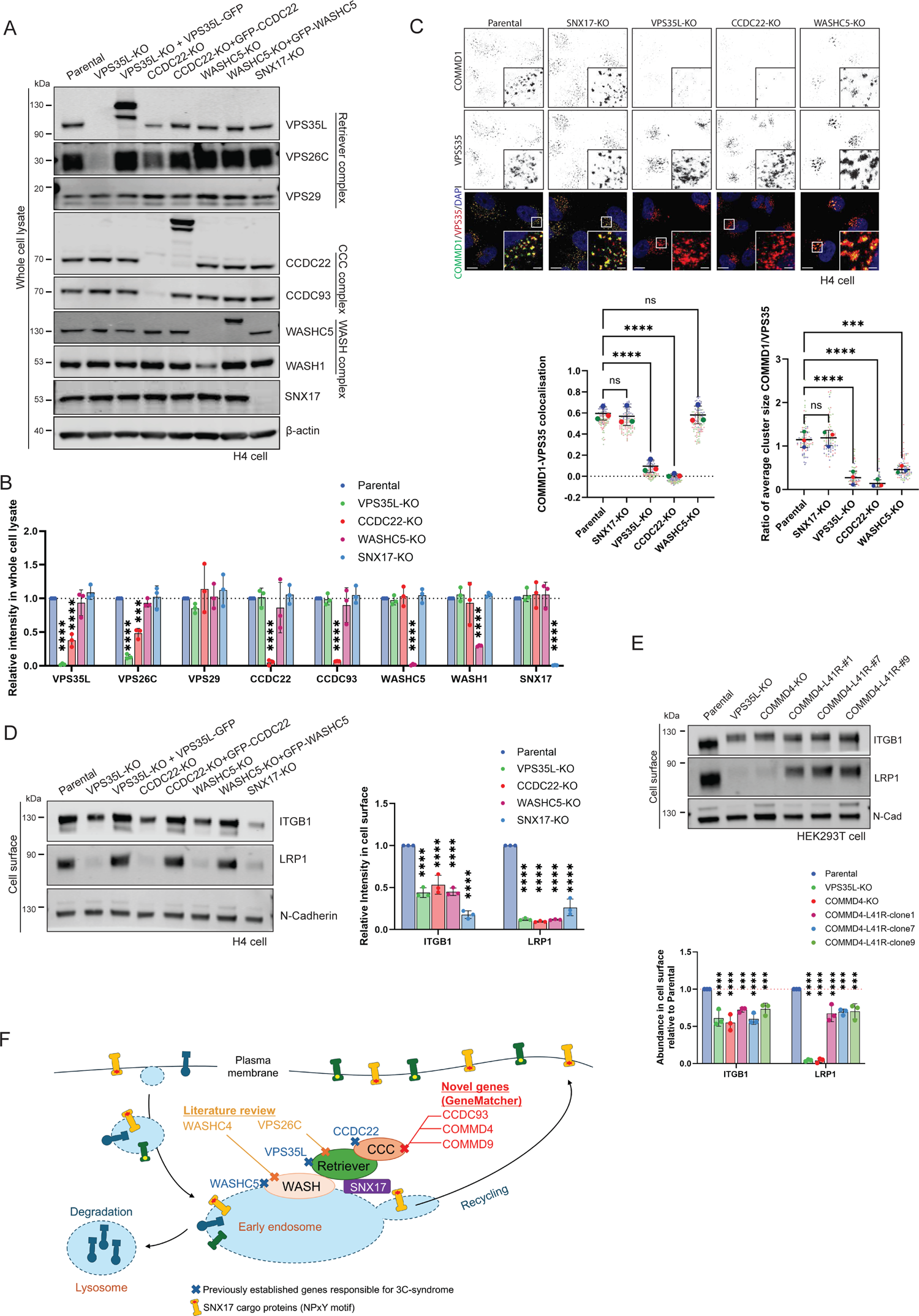
Loss of Retriever/CCC/WASH function disrupts SNX17-cargo recycling (A and B) Representative blots of H4 parental and KO/rescue for VPS35L, CCDC22, WASHC5, and SNX17 using whole cell lysate. β-actin was used as loading control. Bar graphs show relative protein abundance (n=3). (C) Representative view of Immunofluorescence staining of endogenous COMMD1 and VPS35 in H4 cell lines. Graph shows quantification of Pearson’s correlation score from three independent experiments. Pearson’s coefficients for individual cells and means are presented by smaller and larger circles, respectively. Cell numbers analyzed across experiments were 99 parental, 104 SNX17-KO, 99 VPS35L-KO, 108 CCDC22-KO, 135 WASHC5-KO cells. (D) Representative blots of H4 parental and KO/rescue for VPS35L, CCDC22, WASHC5, and SNX17 using cell surface fraction. N-Cadherin was used as loading control. Bar graphs show relative protein abundance (n=3). (E) Representative blots of HEK293T parental, VPS35L-KO, COMMD4-KO, and COMMD4-L41R knock-in cells. N-Cadherin was used as loading control. Bar graphs show relative protein abundance (n=3). (F) Schematic illustration to show accumulation of responsible genes for 3C/Ritscher-Schinzel syndrome in SNX17-Retriever/CCC/WASH recycling pathway. (B-E) Error bars represent mean± SD. **, P<0.01; ****, P<0.0001.

**Figure S3.**
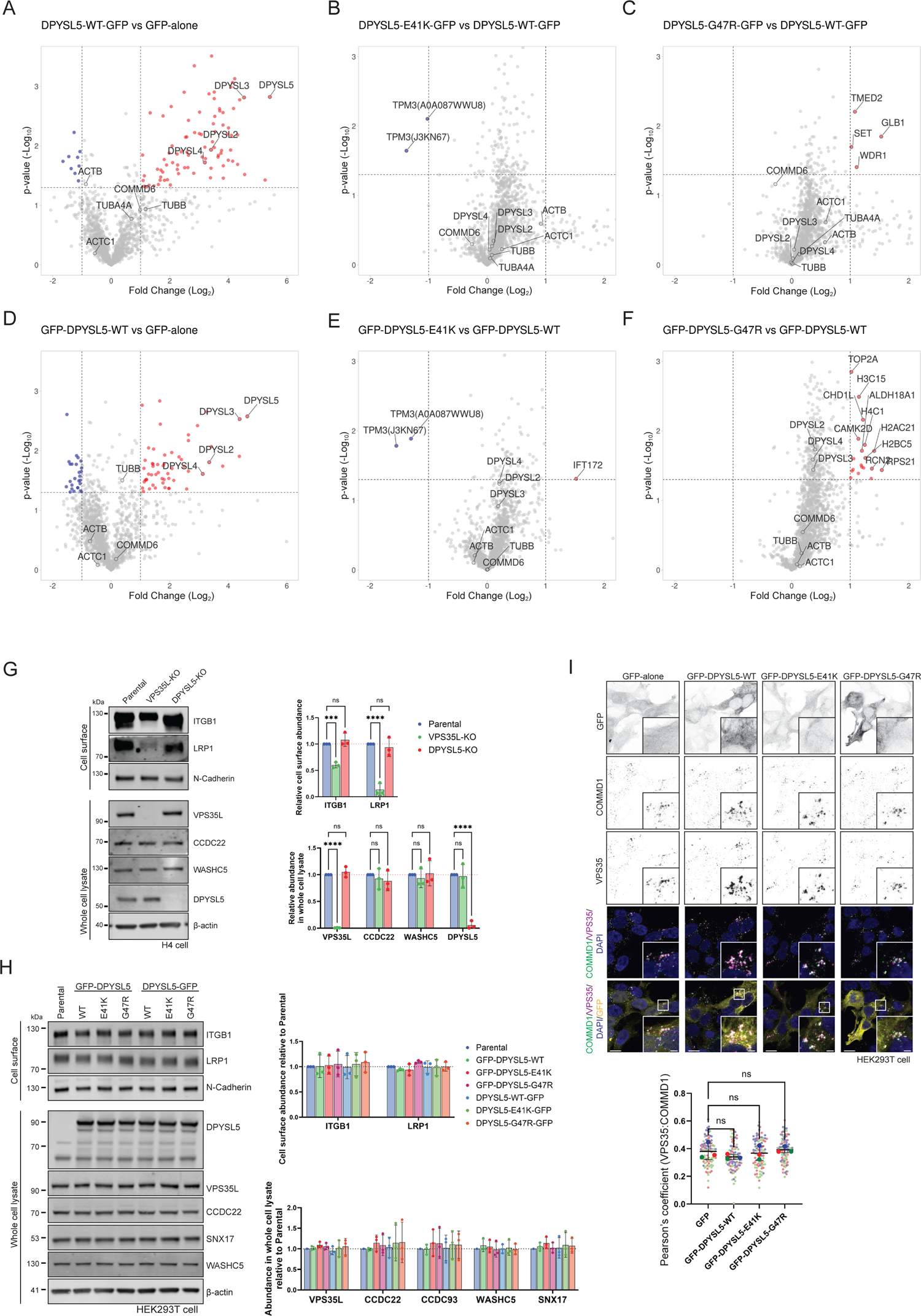
DPYSL5 patient mutants did not affect endosomal recycling (A and B) Volcano plots illustrate enriched (red circles) or depleted (blue circles) interactors in TMT-based proteomics. Wild-type (WT) DPYSL5, DPYSL5-E41K, DPYSL5-G47R, tagged with GFP at either N-terminal or C-terminal end, or GFP-alone were expressed in HEK293T cells, followed by GFP-nanotrap immunoisolation. Comparisons between DPYSL5-WT and GFP-alone are depicted in (A and D), while comparison between GFP-DPYSL5-E41K and GFP-DPYSL5-WT, as well as GFP-DPYSL5-G47R and GFP-DPYSL5-WT, are shown in (B and E) and (C and F), respectively. (G) Representative blots of H4 parental and knock-out of DPYSL5. N-Cadherin and β-actin were used as loading control. Bar graphs show relative protein abundance (n=3). (H) Representative blots of HEK293T parental cell and cells transiently expressing WT-, E41K-, and G47R-DPYSL5 tagged with GFP at either N- or C-terminal end from three independent experiments. N-Cadherin and β-actin were used as loading control. Bar graphs show relative protein abundance. (I) Representative view of Immunofluorescence staining of endogenous COMMD1 and VPS35 in HEK293T cell lines. Scale bars, 10 µm and 2 µm, respectively. Quantification of Pearson’s correlation score from three independent experiments. Cell numbers analyzed across experiments were 96 GFP-alone, 92 GFP-DPYSL5-WT, 91 GFP-DPYSL5-E41K, and 91 GFP-DPYSL5-G47R. Only the cells expressing GFP were taken into account in this analysis. (G-I) Error bars represent mean ± SD.

**Figure S4.**
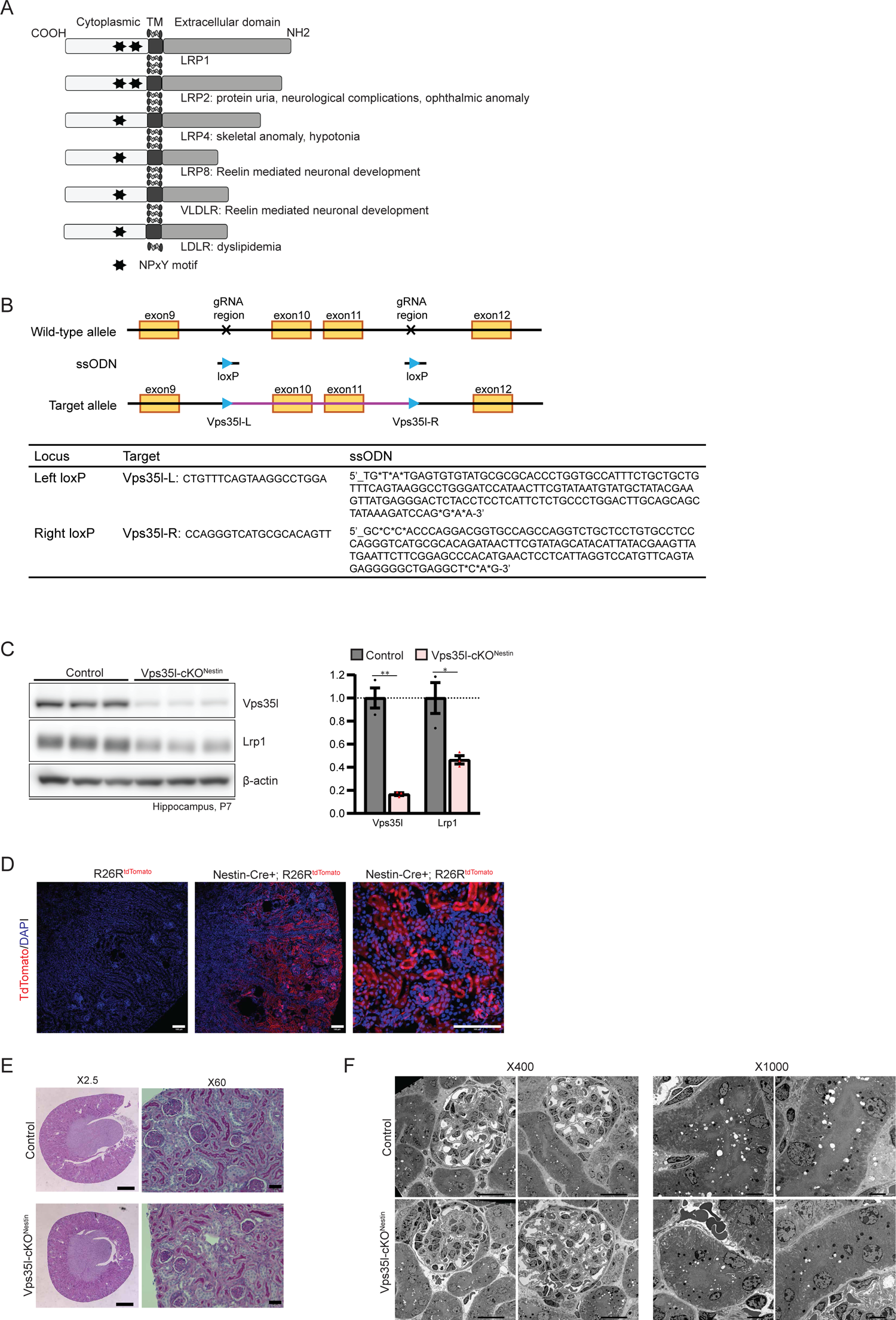
LRP family proteins and generation of VPS35L-flox mouse (A) Schematic illustration of LRP family proteins with NPxY motif in cytoplasmic tail along with linkage to disease phenotypes in loss of their protein function. (B) Generation of Vps35l flox mice. Deletion of exon 10 and 11 induces frame shift which should result in nonsense mediated mRNA decay in the floxed allele when Cre drives recombination. (C) Protein expression of Vps35l and Lrp1 in hippocampal homogenates of control and Vps35l-cKO^Nestin^ mice at P7 (n=3). Bar graph shows band intensities relative to control. Error bars represent mean± SD. *, P<0.05, **, P<0.01. (D) Nestin-Cre;R26R-Tomato double transgenic mice were used to confirm Cre activity in proximal renal tubule. Scale bars, 100 µm. (E) HE staining and weight of the kidney in control and Vps35l-cKO^Nestin^ mice. There were no significant morphological differences between cKO and control mice, although the weight was smaller (n=3). Scale bars, 500 µm and 30 µm, respectively. (F) There was no significant differences in electron microscopy images from Vps35l-cKO^Nestin^ compared to control (n=3). Scale bars, 20 µm and 5 µm, respectively.

**Figure S5.**
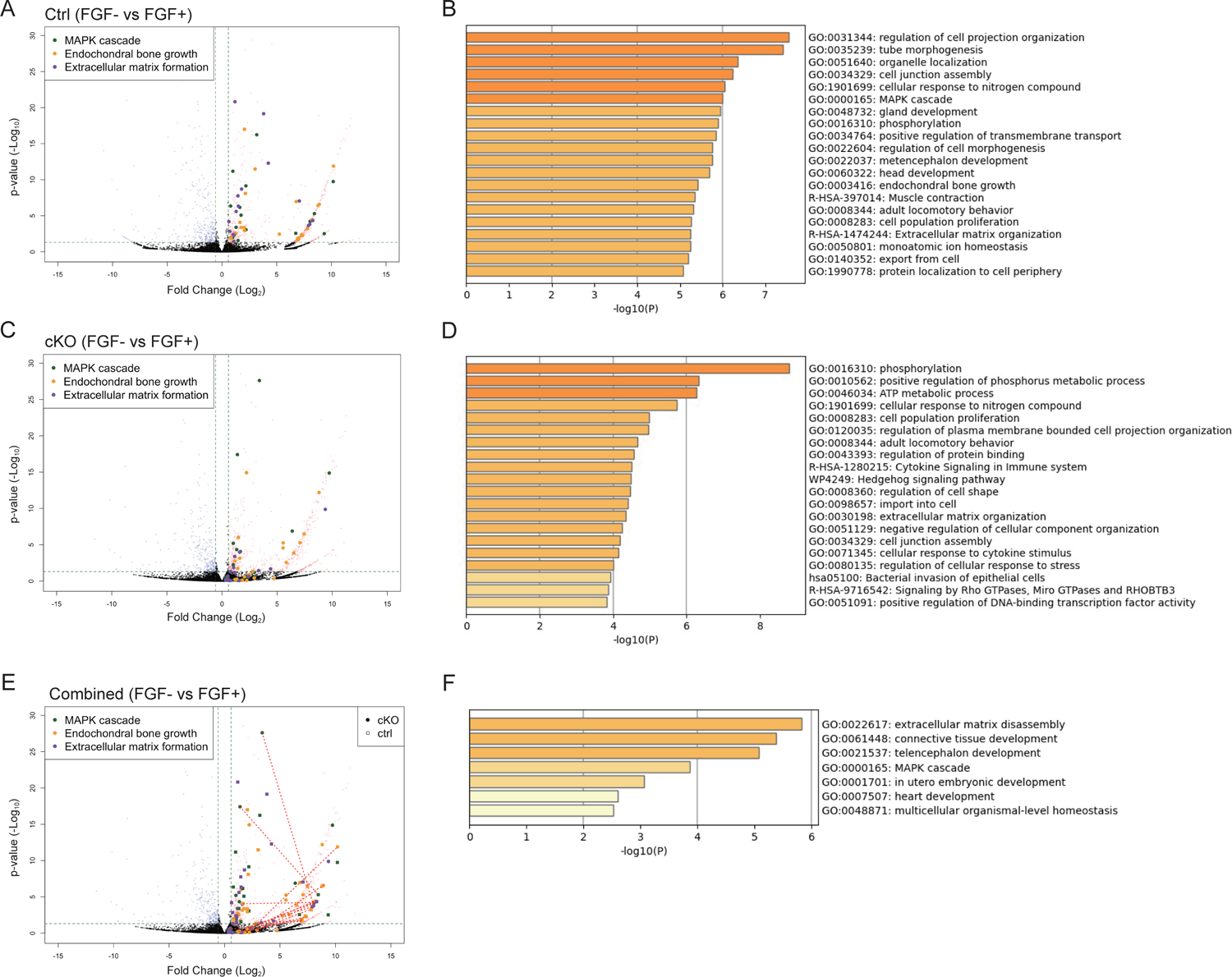
RNA sequencing analysis for ex-vivo cultured chondrocyte showed attenuated MAPK signalling (A-D) Volcano plots and gene enrichment analysis of differential gene expression analysis using RNA-sequencing to compare between samples with and without FGF2 in culture medium. Genes with “MAPK cascade” GO term are labeled with green in volcano blots. Genes with Fold change > 0.585 and q < 0.05 are considered as significant. The results of littermate controls are shown in (A and B), while those of Vps35l-cKO^Prx1^ are shown in (C and D). Both results of control and cKO are blotted together in (E), with the same gene connected by lines to facilitate clear visualization of changes between both genotypes. (F) Gene enrichment analysis of significantly upregulated genes in littermate control compared to Vps35l-cKO^Prx1^.

**Figure S6.**
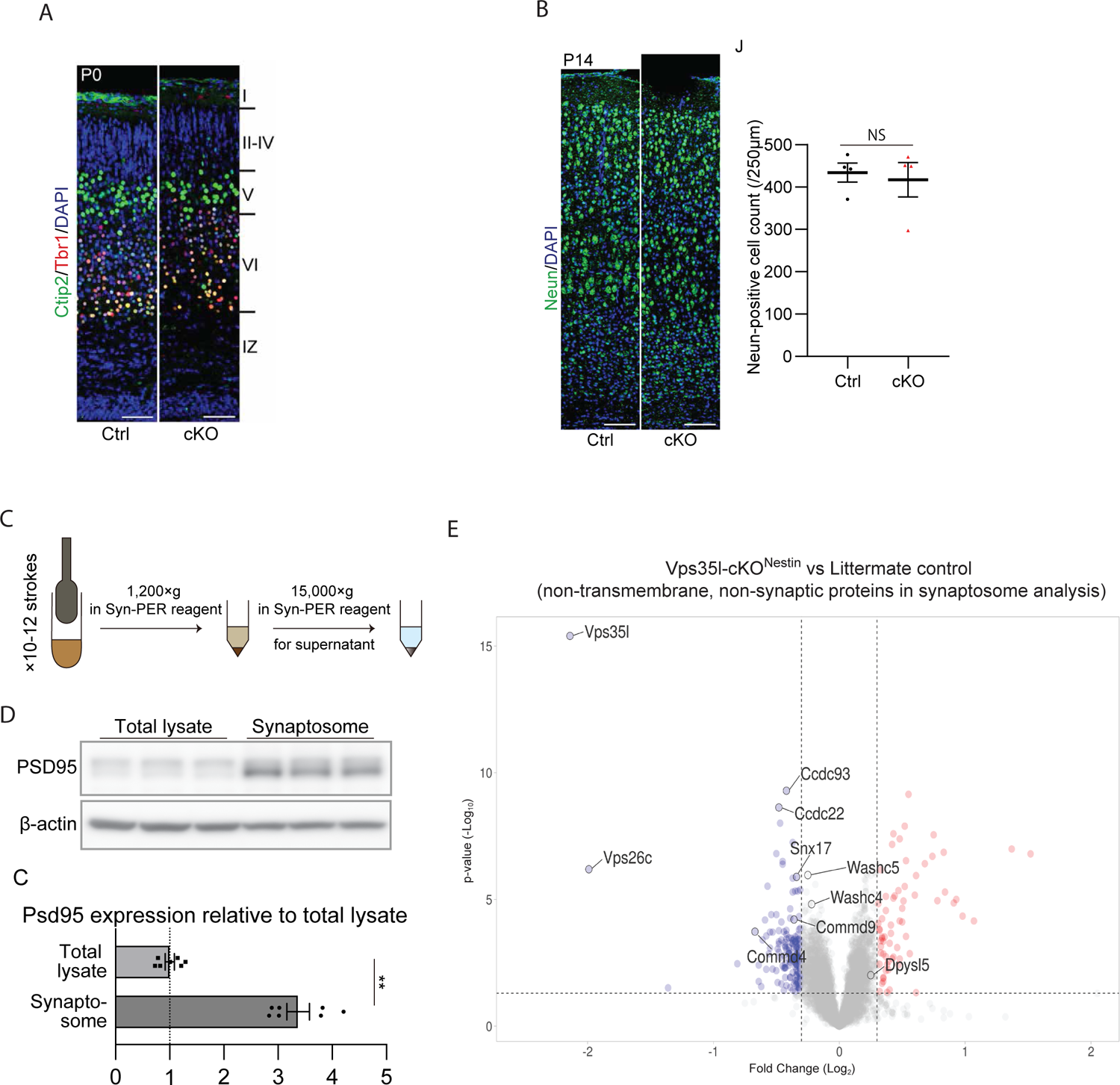
Morphological and synaptosome analysis of VPS35L^Nestin^-cKO mice (A) Immunohistochemistry at P0 of CTIP2 and TBR1 was performed to evaluate neuronal migration. CTIP2 and TBR1 served as markers representing layers V and VI, respectively. [Control; n=4, Vps35l-cKO^Nestin^; n=4]. (B) Immunohistochemistry at P14 of NeuN was performed to evaluate number of neurons. [Control; n=4, Vps35l-cKO^Nestin^; n=4]. (C) Schematic illustration of crude synaptosome isolation. Synaptic fractions were isolated from hippocampal neurons at P7. (D) Western blot analysis for PSD95 confirmed enrichment of synaptic fraction (n=3). (E) Volcano plots of non-transmembrane and non-synaptic proteins with decreased (blue circles) or increased (red circles) abundance in Vps35l-cKO^Nestin^ mice compared to their litter mate controls. Transmembrane and synaptic proteins were plotted in Figure 7G. Synaptic protein fraction were isolated from hippocampal neurons of P7 mice, followed by TMT-based proteome analysis. [Control; n=8, Vps35l-cKO^Nestin^; n=6]. (B and D) Error bars represent mean± SD. **, P<0.01.

**Figure S7.**
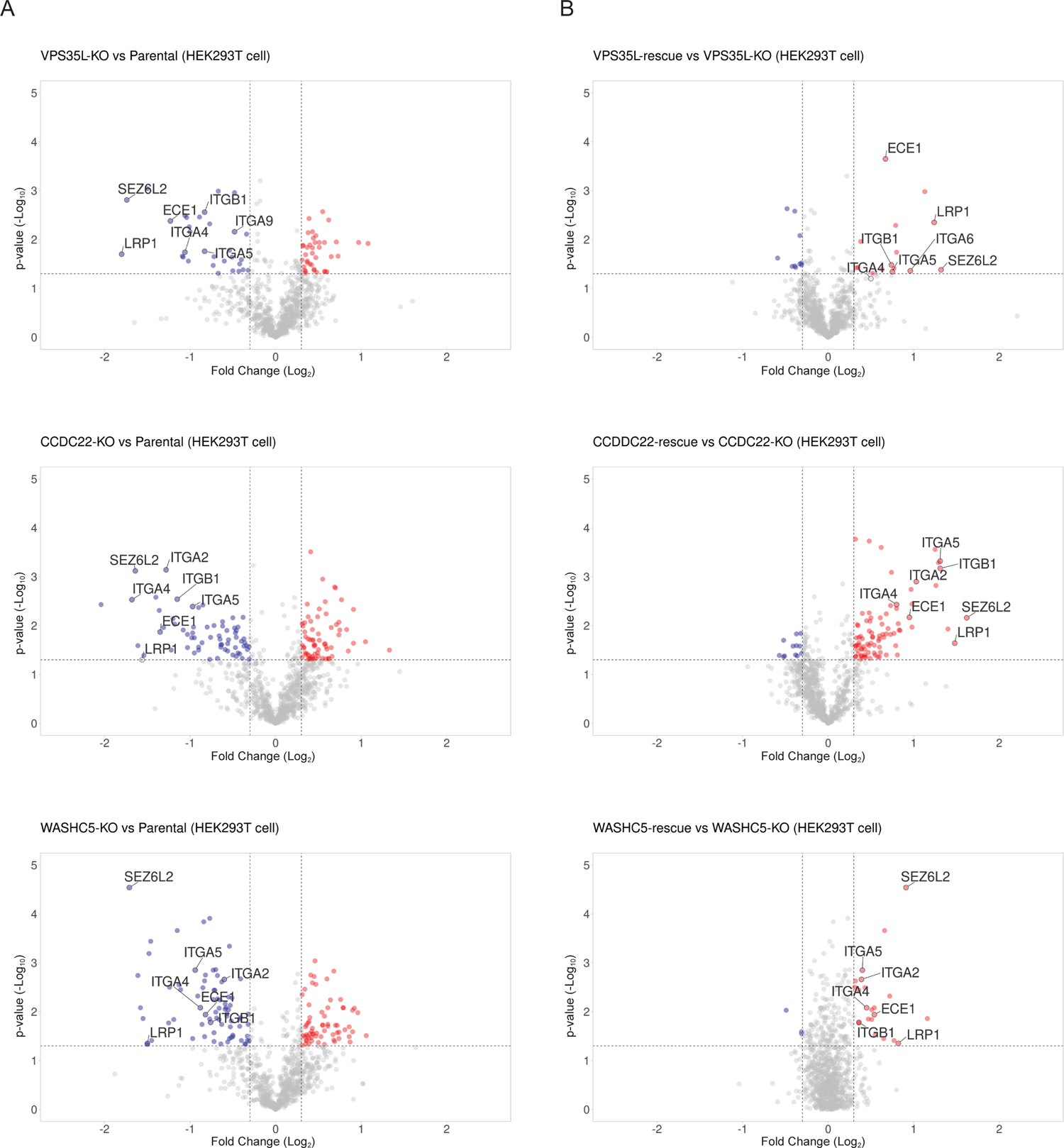
Cell surface proteome analysis in HEK293T cell identified SEZ6L2 and SNX17-cargo proteins affected in VPS35L, CCDC22, and WASHC5 knock-out. (A) Volcano plots of proteins with decreased (blue circles) or increased (red circles) cell surface abundance in knock-out HEK293T cell lines of VPS35L, CCDC22, and WASHC5, from four independent TMT-based proteomics experiments. (B) Volcano plots of proteins with decreased (blue circles) or increased (red circles) cell surface abundance in HEK293T wild-type rescue compared to their knock-out of VPS35L, CCDC22, and WASHC5, from four independent TMT-based proteomics experiments.

**Figure S8.**
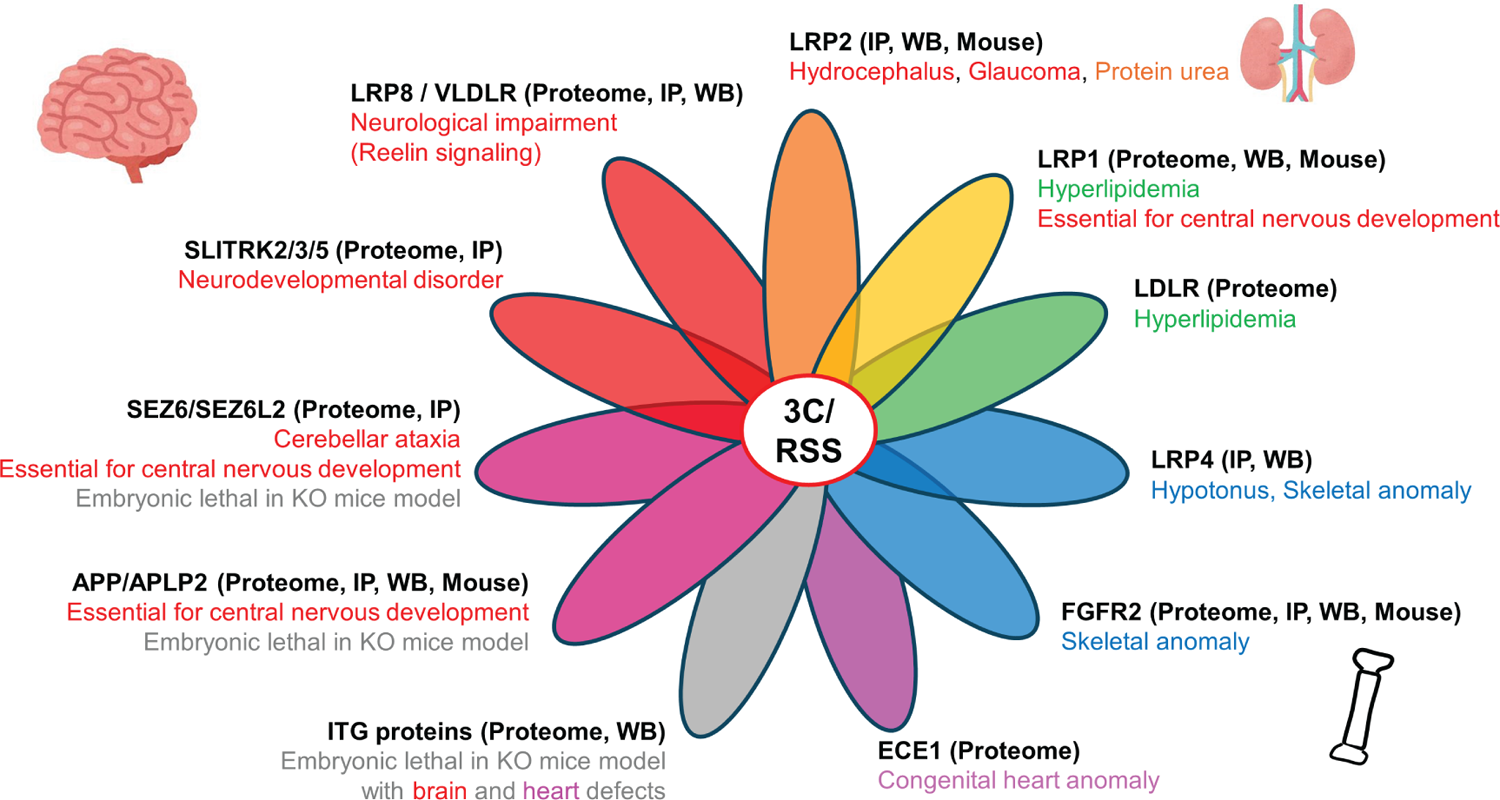
Membrane proteins suggested to be involved in development of clinical phenotypes in Ritscher-Schinzel syndrome. Schematic illustrations showing membrane proteins identified in this study, which are likely responsible for the development of various clinical manifestations in 3C/Ritscher-Schinzel syndrome. IP; Interaction between SNX17 and the NxxY/F motif in the indicated protein was confirmed. WB; Loss of cell surface abundance of the indicated proteins were confirmed by western blotting. Proteome; Loss of cell surface abundance of the indicated proteins were confirmed by proteome analysis. Mouse; Dysregulation of the indicated protein function was confirmed in Vps35l conditional knockout mouse model.

## Supplementary Movie Files

**Movie File 1 and 2**. Tail suspension test of Vps35l-cKO^Nestin^ (Movie File 1) and its littermate control (Movie File 2) at postnatal day 14 (P14). Both mice are mobile, and their hind limbs reflexed but Vps35l-cKO^Nestin^ mice exhibit hindlimb tremor.

**Movie File 3 and 4**. Gait observation of Vps35l-cKO^Nestin^ (Movie File 3) and its littermate control (Movie File 4) at postnatal day 14 (P14). Vps35l-cKO^Nestin^ mice exhibit gait instability as well as tremulous movement.

## Acknowledgements

The authors thank the patients and their parents for participating in this study. We are grateful to Nanako Sugita (Niigata University, Japan) and Ryusaku Esaki (Nagoya University) for technical assistance; Masahiro Yamaguchi (Kochi Medical School, Japan), Rudiger Klein (Max Planck Institute) for materials. We thank the FACS Facility, Wolfson Bioimaging Facility and Proteomics Facility funded by the Wellcome Trust (202904/Z/16/Z and 206181/Z/17/Z) and BBSRC (BB/R000484/1) at the University of Bristol. P.J.C. is supported by the Wellcome Trust (104568/Z/14/Z and 220260/Z/20/Z), the MRC (MR/L007363/1 and MR/P018807/1), the Lister Institute of Preventive Medicine, and the Royal Society Noreen Murray Research Professorship (RSRP/R1/211004). K.K is supported by JSPS KAKENHI (JP20K16897/JP23KJ1817/ JP23K14945/JP23KK0284), Japan Intractable Diseases (Nanbyo) Research Foundation, and Takeda Science Foundation. S.S. is supported by JSPS KAKENHI (JP20K21583), AMED under Grant Number (JP24ek0109646). I.H. is supported by the Platform Project for Supporting Drug Discovery and Life Science Research (Basis for Supporting Innovative Drug Discovery and Life Science Research (BINDS)) from AMED under Grant Number JP21am0101120 (Support No. 2754). K.A.F. is supported by Qatar National Research Fund (QNRF) - NPRP11S-0110-180250. K.S is supported by JSPS KAKENHI (JP20H05700).

## Author contributions

K.K. and P.J.C. designed the research; K.K., K.J.M, E.J., S.Shaw., M.H., R.B., C.D., and K.A.W. performed the biochemistry and cell-based analysis; A.A-M., H.Y.K., M.S.A-H., W.A., A.A.B., J.A., A.S.A.A., E.vB., E.J.M.J., Y.K., M.A.Z. and K.A.F. performed the genetic and clinical analysis of patients; K.K, Y.N., A.Y., S.O., Y.M., H.O., R.K., T.H., I.H., A.S., T.T. and K.S. performed the mouse analysis. P.A.L. and K.J.H. performed the proteomics and bioinformatic analysis; K.K., M.S.Z., K.A.F., S.Saitoh. and P.J.C. acquired funding; K.K. and P.J.C. wrote the 1^st^ draft with all authors contributing to the final manuscript.

## Inclusion and diversity

We support inclusive, diverse, and equitable conduct of research.

## METHODS

### Genetics analysis

Genetic and functional analysis using samples from study participants was approved by the institutional review boards at Nagoya City University (Approval Number: 70-00-0200), Sidra Medicine (Approval Number: IRB 1636872), and National Research Centre, Egypt (Approval Number: 20105). Three unrelated families were recruited via GeneMatcher^30^. Exome and genome sequencing was performed by their local genome diagnostic laboratory and processed according to standardized diagnostic pipelines^72–74^. Written informed consent was obtained from the parents.

### Antibodies

The following antibodies were used in this study (WB: western blot, IF: immunofluorescence): rabbit anti-SNX17 (Proteintech, 10275-1-AP, WB), mouse anti-GFP (Roche, 11814460001, WB), rabbit anti-GFP (GeneTex, GTX30738, WB), mouse anti-mCherry (antibodies.com, A85305, WB), rabbit anti-mCherry (antibodies.com, A85306, WB), rabbit anti-CCDC22, (Proteintech, 16636-1-AP, WB), mouse anti-CCDC93, (Origene, CF800568, WB), rabbit anti COMMD4 and rabbit anti COMMD9 (kind gift from Prof. Ezra Burstein, WB) rabbit anti-C16orf62 (Abcam, ab97889, WB), rabbit anti-C16orf62 (Pierce, PA5-28553, IF), rabbit anti-DSCR3 (Merck Millipore, ABN87, WB), rabbit anti-integrin-β1 (Abcam, ab52971, WB), goat anti-VPS35 (antibodies.com, A83699, IF), mouse anti-VPS29 (Santa Cruz, sc-398874, WB), rabbit anti-KIAA1033 (Proteintech, 51101-1-AP, WB), mouse anti-Strumpellin (Santa Cruz, sc-377146, WB), mouse anti-β actin (Sigma, A1978, WB), rabbit anti-LRP1 (Abcam, ab92544, WB), rabbit anti-LRP2 (Proteintech, 19700-1-AP, WB), rabbit anti-LRP2 (prepared as described in^75^, IHC), rabbit anti-LRP4 (SIGMA, HPA012300, WB), rabbit anti-LRP8 (Abcam, ab108208, WB), mouse anti-VLDLR (Santa Cruz, sc-18824, WB), rabbit anti-APP (Abcam, ab32136, WB), rabbit anti-APLP2 (Proteintech, 15041-1-AP, WB), mouse anti-AP (Thermo Fisher, MA1-20245, WB), rabbit anti-Dab1 (kind gift from Dr. M Hattori, IP/WB)^69^, mouse anti-Phosphotyrosine (Merck, 05-321, WB), rabbit anti-GLUT1 (Abcam, ab115730, IF/WB), mouse anti-N-cadherin (Cell signalling technology, 14215S, WB), anti-PSD95 (Merck, MAB1596, WB), mouse anti-FLAG (SIGMA, F1804, WB), rabbit anti-FGFR2, (Proteintech, 13042-1-AP, WB), rabbit anti-ERK1/2 (Cell Signaling Technology, 9102, WB), rabbit anti-phospho-ERK1/2 (Cell Signaling Technology, 9101, WB), anti Ctip2 (Abcam, ab18465, IHC), anti Tbr1 (Abcam, ab275960, IHC), anti Neun (Cell signalling technology, 12943, IHC). For Odyssey detection of western blots, the following secondary antibodies were used; donkey anti-mouse 680 (Life Technologies), donkey anti-rabbit 800 (Life Technologies).

### Plasmids

Cytoplasmic domain of LRP2, LRP4, LRP8, VLDLR, SLITRK2, SLITRK3, SLITRK4, SLITRK5, APP, APLP1, APLP2, SEZ6, SEZ6L, SEZ6L2 and FGFR2 were cloned from HEK293T DNA or cDNA into pEGFPC1 vector which contains signal peptide cloned from CI-MPR as previously described^76^) The expression plasmid for AP-RR36 to express central fragment of Reelin (RR3 to RR6) and whole length of human Reelin were previously described^69,77^.

### Lentivirus production

For lentivirus production, shRNAs driven by a H1 promoter were generated for the knockdown of rat SNX17 (target sequence #1: 5’-gtacatgcaagctgttcgg-3’, #2: 5’-gatcgtgctcagaaagagt-3’), rat VPS35L (target sequence #1: 5’-caccagtgttattcagttcta-3’, #4: 5’-tacggagaagctgtctattaa-3’), mouse SNX17 (target sequence #1: 5’-gtacatgcaagctgttcgg-3’, #2: 5’-gattgtgctcagaaagagt-3’), mouse VPS35L (target sequence #1: 5’-cacgagtgttattcagttcta-3’, #2: 5’-cacgagtgttattcagttcta-3’), and a scramble control (non-targeting sequence: 5’-aattctccgaacgtgtcac-3’). Oligonucleotides were cloned into a modified pXLG3-GFP viral vector and co-transfected into a 15 cm dish of HEK293T cells with the helper plasmids Pax2/p8.91 and pMDG2 using PEI. For primary culture, the viruses were harvested 48 hr after transfection, spin down at 4000 rpm for 10 min at room temperature (RT) and filtered through 0.45 mm filters.

### Cell Culture and transfection

HEK293T and MC3T3-E1 (subclone 4) cell lines were originally sourced from American Type Culture Collection, and H4 cell line was a gift from Dr Helen Scott and Professor James Uney. Cells were grown in DMEM medium (Sigma-Aldrich) for HEK293T and H4 or MEM-α(Thermo Fisher SCIENTIFIC, A1049001) for MC3T3-E1 supplemented with 10% (vol/vol) FBS (Sigma-Aldrich) and penicillin/streptomycin (Gibco), and grown under standard conditions. All tissue culturing materials were sterile and tissue culture was performed in Class II vertical laminar flow cabinets.

FuGENE HD (Promega) was used for transient transfection of DNA according to the manufacturer’s instructions. For the isolation of CRISPR–Cas9 clonal lines, HEK293T or H4 cells were transfected with pX459 plasmid coding for the genomic RNA (gRNA) of the gene of interest. The cells were then subjected to puromycin selection for 24 - 48h. The cells were subsequently resuspended and plated for single cell isolation in Iscov’s modified Dulbecco’s medium (Sigma-Aldrich) supplemented with 10% (vol/vol) FBS (Sigma-Aldrich) penicillin/streptomycin (Gibco). Once the cell colonies were expanded, they were subjected to lysis and western blotting to determine the levels of the target proteins. The VPS35L and VPS35 knockout cell lines were characterized previously^18,78^.

Primary neuronal cultures were prepared from embryonic day E18 Wistar rat brains as previously described (Martin and Henley, 2004). In brief, dissociated cortical cells were grown in six well dishes (500,000 cells/well) coated with poly-L-lysine (P2636; Sigma-Aldrich) in 2 ml plating medium (Neurobasal medium (21103–049, Gibco) supplemented with 5% horse serum (H1270), 2% B27 (17504–044, Gibco) and 1% Glutamax (35050–038)) which was exchanged for 2 ml feeding medium 2 hr after plating (Neuro basal medium (21103–049, Gibco), 2% B27 (17504–044, Gibco), and 0.4% Glutamax (35050–038)). Cells were then fed with an additional 1 ml of feeding medium 7 days after plating. Neurons were transduced with shRNA viruses on between DIV12 and DIV14 then left for 5 days before analysis. MC3T3E1 cells were transduced with shRNA viruses and incubated for 24 h to allow viral infection, then cultured with fresh complete medium for another 24 h, allowing cell recovery and shRNA expression. After 48 h from viral infection, puromycin was added to select cells stably expressing shRNA. Cells were used within eight passages from the purchased stock.

### CRISPR/Cas9 mediated gene editing

All the CRISPR/Cas9 target sequences were designed by CRISPRdirect^79^. 5’-GGGTCACCCGCATGCGATGC<u>TGG</u> for SNX17, 5’-TTGTGTATTACACTGTGCGA<u>GGG</u> for LRP2, 5’-TGATCACAATGCAGATCGTT<u>GGG</u> for WASHC5, 5’-<u>CCT</u>CGCTCCTCGAACACCATGCC for CCDC22, 5’-CCTGGCAGAAATCAGCACGC<u>TGG</u> for COMMD4, 5’-<u>CCT</u>CGAAAGATGTTGTCAGACAG for COMMD9, 5’-TTTCAAGAAAACTCTACGAT<u>AGG</u> for CCDC93, 5’-CGTGGTGGTCATATCGAGTA<u>AGG</u> for VPS26C, 5’-GAGAAGAATTGAGGACGCTC<u>TGG</u> for WASHC4, 5’-<u>CCC</u>ACGAGGCTGACGTCTACATC for DPYSL5, which were cloned into px459 vector.^80^ For generating COMMD4-L41R point mutation in HEK293T cell line, we used CRISPR/Cas9 mediated homology-directed repair (HDR) pathway. Target sequence was 5’-CCAGGTACTAAAGGAGC[T>G]GC<u>TGG,</u> and we used single-stranded DNA donor with T>G substitution mimicking patient-derived L41R mutation as template. ssODN sequence was as followed:GTGAAGTTGCGGCTGCTCTGCAGCCAGGTACTAAAGGAGC<u>G</u>GCT<u>C</u>GGACAGGGGATTGATGTGAGTACAAGATCCAGCACC. We employed silent mutation (G>C) on the template ssODN to disrupt PAM sequence. Clones were isolated and gene disruption or point mutation were validated by PCR and PCR-based sequencing.

### Generation of stable lentiviral HEK293T and H4 cell lines

The gene of interest was subcloned into the lentiviral vector pXLG3 or pLVX for the generation of lentiviral particles. Lentiviral particles were produced and harvested in HEK293T cells. Transduced cells were grown in DMEM supplemented with 10% (vol/vol) FBS and penicillin/streptomycin and grown at 37°C in a 5% CO2 incubator.

### Surface biotinylations

All solutions were pre-chilled to 4°C and all steps were carried out on ice to prevent internalisation. Fresh membrane impermeable Sulpho NHS-SS-Biotin (21331, Thermo Fisher Scientific) was dissolved in PBS at a final concentration of 0.2 mg/ml. Cells were washed twice in ice-cold PBS before being incubated with biotin for 30 mins at 4 °C. The cells were then washed in PBS before being quenched in quenching buffer (50 mM Triz, 100 mM NaCl, pH 7.5) for 10 min at 4°C. The cells were then lysed in 2% Triton-X-100 (X100, Sigma) plus protease inhibitor cocktail tablets (A32953, Thermo Fisher Scientific) in PBS, and subjected to Streptavidin-bead-based affinity isolation (GE Healthcare).

### Immunoprecipitation and western blot analysis

HEK293T cells transfected with GFP-, mCherry-, FLAG-tagged protein constructs were lysed 24 or 48 h post transfection. The cells were washed with PBS, then 0.5 ml lysis buffer (20mM HEPES pH7.4, NaCl 150mM, Triton-X 0.5%) was added to 15 cm dishes. The lysates were cleared by centrifugation, and supernatants were incubated with 15 microlitres of GFP-/mCherry-trap beads (Chromotek) or FLAG-trap beads (SIGMA) on a rocker at 4 °C for 30 min. Following incubation, trap beads were pelleted and washed with lysis buffer three times. After the final wash, lysis buffer was removed and beads were resuspended in 2x NuPAGE LDS Sample Buffer (Life Technologies), 5% β-mercaptoethanol and boiled at 95 °C for 5 min. The samples were then loaded for SDS-PAGE and immunoblot analysis.

### TMT labelling and proteome analysis

The TMT analysis of the cell surface proteome samples and the GFP-nanotrap samples has been performed in the same way. i.e. GFP-nanotrap samples were reduced (10mM TCEP, 55°C for 1h), alkylated (18.75mM iodoacetamide, room temperature for 30min.) and then digested from the beads with trypsin (1.25µg trypsin; 37°C, overnight). The resulting peptides were then labeled with TMT ten-plex reagents according to the manufacturer’s protocol (Thermo Fisher Scientific, Loughborough, LE11 5RG, UK) and the labelled samples pooled and desalted using a SepPak cartridge according to the manufacturer’s instructions (Waters, Milford, Massachusetts, USA). Eluate from the SepPak cartridge was evaporated to dryness and resuspended in buffer A (20 mM ammonium hydroxide, pH 10) prior to fractionation by high pH reversed-phase chromatography using an Ultimate 3000 liquid chromatography system (Thermo Scientific). In brief, the sample was loaded onto an XBridge BEH C18 Column (130Å, 3.5 µm, 2.1 mm X 150 mm, Waters, UK) in buffer A and peptides eluted with an increasing gradient of buffer B (20 mM Ammonium Hydroxide in acetonitrile, pH 10) from 0-95% over 60 minutes. The resulting fractions (concatenated into 5 in total) were evaporated to dryness and resuspended in 1% formic acid prior to analysis by nano-LC MSMS using an Orbitrap Fusion Tribrid mass spectrometer (Thermo Scientific).

High pH RP fractions were further fractionated using an Ultimate 3000 nano-LC system in line with an Orbitrap Fusion Tribrid mass spectrometer (Thermo Scientific). In brief, peptides in 1% (vol/vol) formic acid were injected onto an Acclaim PepMap C18 nano-trap column (Thermo Scientific). After washing with 0.5% (vol/vol) acetonitrile 0.1% (vol/vol) formic acid peptides were resolved on a 250 mm × 75 μm Acclaim PepMap C18 reverse phase analytical column (Thermo Scientific) over a 150 min organic gradient, using 7 gradient segments (1-6% solvent B over 1min., 6-15% B over 58min., 15-32%B over 58min., 32-40%B over 5min., 40-90%B over 1min., held at 90%B for 6min and then reduced to 1%B over 1min.) with a flow rate of 300 nl min^−1^. Solvent A was 0.1% formic acid and Solvent B was aqueous 80% acetonitrile in 0.1% formic acid. Peptides were ionized by nano-electrospray ionization at 2.0kV using a stainless-steel emitter with an internal diameter of 30 μm (Thermo Scientific) and a capillary temperature of 275°C.

All spectra were acquired using an Orbitrap Fusion Tribrid mass spectrometer controlled by Xcalibur 2.1 software (Thermo Scientific) and operated in data-dependent acquisition mode using an SPS-MS3 workflow. FTMS1 spectra were collected at a resolution of 120 000, with an automatic gain control (AGC) target of 200 000 and a max injection time of 50ms. Precursors were filtered with an intensity threshold of 5000, according to charge state (to include charge states 2-7) and with monoisotopic peak determination set to peptide. Previously interrogated precursors were excluded using a dynamic window (60s +/-10ppm). The MS2 precursors were isolated with a quadrupole isolation window of 1.2m/z. ITMS2 spectra were collected with an AGC target of 10 000, max injection time of 70ms and CID collision energy of 35%.

For FTMS3 analysis, the Orbitrap was operated at 50 000 resolution with an AGC target of 50 000 and a max injection time of 105ms. Precursors were fragmented by high energy collision dissociation (HCD) at a normalised collision energy of 60% to ensure maximal TMT reporter ion yield. Synchronous Precursor Selection (SPS) was enabled to include up to 10 MS2 fragment ions in the FTMS3 scan.

### MS data analysis

The MS data were searched against the human Uniprot database. Protein groupings were determined by Proteome Discoverer software v2.4 (Thermo Scientific), however, the master protein selection was improved with an in-house script. The script first searches Uniprot for the current status of all protein accessions and updates redirected or obsolete accessions. The script further takes the candidate master proteins for each group, and uses current uniprot review and annotation status to select the best annotated protein as master protein without loss of identification or quantification quality. The protein abundances for each sample were normalised such that all samples had an equal total protein abundance, then normalised abundances were Log2 transformed to bring them closer to a normal distribution. Univariate paired t-tests or linear model analysis were performed for each comparison of interest using normalised abundances. For all comparisons, the p-value was adjusted for multiple testing using the Benjamini-Hochberg method.

### Immunofluorescence staining

Cells on coverslips were fixed in 4% paraformaldehyde (PFA) for 20min at room temperature or methanol at −20 °C for 7min. Cells fixed with PFA were then permeabilized with 0.1% Saponin (SIGMA) for 5min. The fixed cells were blocked in 2% BSA for 30 min followed by incubation with primary antibodies for 1 h and fluorophore-conjugated secondary antibodies (Alexa Fluor; Thermo Fisher Scientific) for 30 min Following the incubation, coverslips were washed three times in PBS, then mounted with Fluoromount (Sigma) onto glass slides.

### Reelin uptake and Dab1 phosphorylation assay

Primary rat cortical neurons were transduced with shRNA viruses on DIV12 for five days before analysis. Conditioned medium containing Reelin-containing or AP-Reelin-RR36-containing was prepared as previously described.^69^ Briefly, expression plasmid was transfected into HEK293T cells using PEI followed by replacement of medium with neuronal feeding medium, then cells were incubated for 48 h. For Reelin uptake assay, cells were incubated in AP-RR36 conditioned medium for 30 min, then cells were washed with ice-cold PBS for three time and lysed with lysis buffer. Samples were analysed by SDS-PAGE and immunoblot analysis. Induced phosphorylation of Dab1 in rat cortical neurons was measured essentially as previously described^77^. Briefly, cortical neuronal cells were incubated with Reelin-containing media for 20 min at 37 °C, then lysed, and endogenous immunoprecipitation was performed with anti-Dab1 antibody. Samples were analysed by Western blotting using anti-Dab1 and anti-phosphotyrosine antibody to quantify phosphorylation status of Dab1.

### mCherry-D3 uptake assay

The construct to express mCherry-tagged D3 fragment of RAP (Receptor associated protein) was kindly provided from Prof. Martin Lowe. mCherry-D3 was prepared as described previously.^81^ In brief, mCherry-D3 was expressed in E. coli BL21 cells by induction with 1 mM IPTG for 16 hours at 18°C. The protein was purified using Ni-NTA agarose using standard methods, snap frozen in liquid nitrogen, and stored at −80°C until use. Uptake experiments were performed with HEK293T cells, where cells were incubated with purified mCherry-D3 in complete medium for 30 min, then uptake was monitored by lysing the cells followed by SDS-PAGE and immunoblot analysis.

### FGF2 uptake assay

MC3T3-E1 cells were cultured with serum free MEM-αmedium with 5ng/ml Human FGF-2 recombinant protein for overnight, then medium was replaced with one containing 100ng/ml of FGF-2 and incubated for 7 min. Cells were lysed and subjected to SDS-PAGE and immunoblot analysis.

### Animals

C57BL/6 mice (CLEA Japan), B6D2F1 (CLEA Japan) and ICR mice (Charles River Japan) were used. Nestin-Cre mice were obtained from Dr. Rudiger Klein (Max Planck Institute)^82,83^. Rosa-tdTomato reporter mice (B6. Cg-Gt(ROSA)26Sortm14(CAG-tdTomato)Hze/J; Stock no. 7914; The Jackson Laboratory) were provided by Dr. Masahiro Yamaguchi (Kochi Medical School, Kochi, Japan). Prx1-Cre mice (B6.Cg-Tg(Prrx1-cre)1Cjt/J; Stock no. 5584) were purchased from the Jackson Laboratory. Vps35l^flox/flox^ mice were generated in this study as described below. All mice used for this study were housed at temperature and humidity ranges of 21–23 °C and 40–60%, respectively, with a 12-h light– dark cycle. All animals were fed food and water adlibitum. All animals were housed in standard polycarbonate cages in groups of same-sex littermates. All animal experiments were conducted according to relevant national and international guidelines contained in the Act on Welfare and Management of Animals (Ministry of Environment of Japan) and the Regulation of Laboratory Animals (Nagoya City University) and under the protocols approved by the Institutional Animal Care and Use committee review panels at Nagoya City University, and by the Animal Care and Experimentation Committee of Gunma University.

### Generation of *Vps35l*^flox/flox^ mice

*Vps35l*^flox/flox^ mice were generated using the sequential electroporation method as previously reported^84,85^. Synthetic crRNA (Alt-R CRISPR–Cas9 crRNA; IDT) and tracrRNA (Alt-R CRISPR–Cas9 tracrRNA, IDT) were used in this study. Corresponding donor single-stranded oligodeoxynucleotides (ssODNs) with 5’- and 3’-homology arms flanking *loxP* and a restriction site were also designed (Figure. S?). Pre-annealed crRNA/tracrRNA (3 μM), recombinant Cas9 protein (100 ng μl^−1^; GeneArt Platinum Cas9 Nuclease, Thermo Fisher Scientific) and ssODN (400 ng μl^−1^; Ultramer, IDT) in Opti-MEM I (Life Technologies) were used for electroporation medium. Fertilized eggs were isolated from a superovulated B6D2F1 female mice 21-h post-administration of human chorionic gonadotropin (hCG). The first electroporation to insert left *loxP* was conducted at the one-cell stage (24–26-h post-hCG); the second electroporation to insert right *loxP* was conducted at the two-cell stage (42–44-h post-hCG). The sequentially electroporated embryos were then transferred to the oviduct of pseudopregnant ICR females. Alleles flanked by *loxP* of obtained mice were confirmed by sequencing of PCR products using the following primer sets: left *loxP*: 5’-TCCCAACTCTGGAAGCAGAT-3’ and 5’-TGTCCCAAACTCTGGCTTTT-3’, right *loxP*: 5’-CTCATCGCCCGTGTGTCT-3’ and 5’-TCACATCTGTGAGAAATGACAGC-3’.

### Dissection of embryonic tibiae and *Ex vivo* culture

Ex vivo culture of tibiae was performed as previously described^86^. In brief, the tibiae of VPS35L^Prx1^ KO mice and littermate controls (C57BL/6 background) were anesthetize with isoflurane and dissected under a microscope on embryonic day 16.5. Thereafter, tibiae were cultured in a 24-well plate with α-minimal essential medium supplemented with 0.2% bovine serum albumin, 1 mM β-glycerophosphate, 50 µg/ml ascorbic acid, 1% penicillin-streptomycin. The tibiae from the same individual were separated into two groups, one for FGF2+ cultured with FGF2 (3339-FB-025, R and D Systems) and the other for FGF2-cultured without FGF2 for 4 days. The medium was changed daily.

### RNA extraction and sequencing

The tibiae with or without *ex vivo* culture was dissociated with 0.3% Collagenase A (Roche) in DMEM at 37 °C for 1.5 hours and pipetting in and out to remove the connective tissues surrounding the tibiae. Then, the tibiae were washed with PBS and incubated again with 0.3% Collagenase A (Roche) in DMEM at 37 °C for 2.5 hours, dissociated by pipetting in and out, and dissociated chondrocytes were collected with centrifugation at 1000 rpm for 3 minutes. Total RNA was extracted using RNeasy Mini Kit (Qiagen) and library preparation was performed with NEBNext® Poly(A) mRNA Magnetic Isolation Module and MGIEasyRNA Directional Library Prep Set, followed by the sequence with DNBSEQ-G400RS (MGI).

### Transcriptome analysis

The quality of the fastq files obtained from the RNA sequence was checked and the adapter sequences were removed using Trim Galore, mapped at reference genome (hg19) with HISAT2, followed by the read count using StringTie. The count data were analyzed with DESeq2 and visualized with R packages.

### Tissue harvest and sectioning

The mice were deeply anesthetized with isoflurane, decapitated, and perfused with PBS followed by 4% paraformaldehyde. The brain was then immersed in 4% paraformaldehyde overnight. For the cryosection, the fixed brains were immersed in 10%, 20%, and 30% sucrose solutions. After replacement with sucrose solutions, organs were embedded and frozen in SCEM (Super Cryoembedding Medium, SECTION-LABCo., Ltd.) and were sectioned at 20-mm thickness for immunohistochemistry using cryomicrotome (CM3050, Leica). For Nissl and hematoxylin and eosin staining, the dissected and post-fixed brains were dehydrated in ethanol and embedded in paraffin, followed by sectioning at 4mm thickness.

### Immunohistochemistry

For immunohistochemistry in brain tissue, cryo-sectioned tissues were incubated in a blocking solution of 0.5% Triton X-100 in Blocking One (Na-calai Tesque, Inc.) at room temperature for 60 min and then incubated with primary antibodies of interest at 4 °C for 16–24 h. Samples were then washed with PBS-T and stained with secondary antibody and DAPI at 4 °C for 2 h. After final washing with PBS, the samples were sealed with mounting medium and visualized by fluorescence microscopy (Nikon A1RS+). For immunohistochemistry in kidney tissue, sections were deparaffinized by immersion in xylene and rehydrated through a graded ethanol series. Antigen retrieval was accomplished by heating the slides in a microwave in 10 mM citrate buffer, pH 6.0. To eliminate endogenous peroxidase activity, the sections were incubated in 3% H2O2. Following incubation with 2.5% normal goat serum to block nonspecific binding, the sections were incubated with anti-megalin antiserum ^75^ in PBS containing 1% goat serum for 60 minutes at room temperature. This step was followed by incubating for 30 minutes with ImmPRESS – HRP Goat Anti-Rabbit IgG Polymer Reagent (Vector Laboratories, Burlingame, CA). Finally, peroxidase activity was visualized with the 3,39-Diaminobenzidine substrate-chromogen system (Dako), counterstained with Mayer haematoxylin, and examined under a light microscope.

### Synaptosome preparation

The mice were deeply anesthetized with isoflurane, decapitated, and perfused with PBS. The hippocampus was harvested under a microscope on postnatal day 7, followed by synaptosome isolation as previously described^65^. Briefly, each hippocampus was transferred to a glass homogenizer and Syn-PER reagent was added to the tissue, followed by the homogenization on ice with slow stokes (∼10 strokes). The homogenate was transferred to a fresh centrifuge tube and centrifuged at 1200 x g for 10 min at 4°C and the supernatant was transferred to a fresh tube and centrifuged again at 15,000 x g for 20 min at 4°C. Then, the pellet was resuspended in Syn-PER reagent and Triton X-100 and SDS were added to a final concentration of 1% and 0.1%, respectively, and left at 4°C on a rotating wheel for 1h to lyse synaptosomes and solubilize membrane proteins. The samples were then centrifuged at 16,000 x g for 20 min and the supernatant was used for proteome analysis.

### Statistical analysis

The Odyssey infrared scanning system (LI-COR Biosciences, USA) was used for quantitation of Western blot analysis. Colocalisation analysis of fluorescently labelled proteins was performed in Volocity 6.3 software (PerkinElmer). All quantified data was shown as the mean ± SD measurement. Two-side Student’s t-test was performed to compare the means between two groups. One-way or Two-way analysis of variance (ANOVA) with post-hoc Dunnett’s honestly significance difference calculator test was used. Statistics were calculated using GraphPad Prism 10 (GraphPad Software, USA).

### Supplementary clinical information

Detailed clinical information was removed due to medRxiv policy. These details are available upon request from the corresponding authors.

**Supplementary Table 1.** Clinical comparison among patients with COMMD4, COMMD9, or CCDC93 mutations.

|  | This study |  |  |  |  |
| --- | --- | --- | --- | --- | --- |
|  | COMMD4-II-3 | COMMD4-II-4 | COMMD4-II-5 | COMMD9 | CCDC93 |
| Identified variants | p.Leu41Arg | p.Leu41Arg | p.Leu41Arg | p.Leu70GlyfsTer5 | p.Ser631Ala, p.Tyr457Ter |
| Inheritance | homozygous | homozygous | homozygous | homozygous | compound heterozygous |
| Gender / age (years) | Female / 0-5* | Male / 0-5* | Female / 0-5* | Male / 0-5 | Male / 0-5 |
| Craniofacial findings |  |  |  |  |  |
| Aplasia cutis on skull | n.a. | n.a. | n.a. | + | + |
| Relative macrocephaly | Microcephaly | n.a. | Microcephaly | + | + |
| Large anterior fontanelle | + | n.a. | + | - | + |
| High and broad forehead | n.a. | n.a. | n.a. | + | + |
| Arched wide eyebrow | n.a. | n.a. | n.a. | + | + |
| Hypertelorism | n.a. | n.a. | n.a. | + | + |
| Thin upper lip | n.a. | n.a. | n.a. | + | + |
| Long philtrum | n.a. | n.a. | n.a. | + | + |
| Ear | Low set ears | n.a. | n.a. | Over folded helix<br>Low set large ears | Over folded helix<br>Low set small ears |
| Postnatal growth retardation | n.a. | n.a. | n.a. | BW -1.4SD<br>Length -2.5SD<br>HC -0.9SD | BW +0.4SD<br>Height -2.8SD<br>HC +1.5SD |
| Developmental delay / ID | n.a. | n.a. | + | + | + |
| Brain image findings |  |  |  |  |  |
| Cerebellum | Inferior vermian<br>hypoplasia | Inferior vermian<br>hypoplasia | Inferior vermian<br>hypoplasia | Vermis hypoplasia | Vermis hypoplasia |
| Other findings | Corpus collosum<br>dysgenesis,<br>Choroid plexus cyst,<br>Widened extra-axial spaces | Corpus collosum<br>dysgenesis,<br>Choroid plexus cyst | Corpus callosum<br>dysgenesis. Choroid<br>plexus cyst | Dysgyria,<br>Periventricular<br>demyelination,<br>Thin corpus callosum | Increased infratentorial<br>space with high position<br>of the tentorium, |
| Cardiac disorder | Small fenestrated ASD | Small Secundum ASD | Multiple small VSD | - | - |
| Limb anomalies | - | n.a. | - | Brachydactyly<br>Reduced distal creases<br>Arthrogryposis<br>Radioulnar dysostosis<br>Overriding toes | Brachydactyly<br>Nail hypoplasia<br>Overlapping toes |
| Ophthalmologic changes | Retinopathy | n.a. | Retinopathy | - | - |
| Proteinuria | n.a. | n.a. | + | - | + |
| Dyslipidemia | n.a. | n.a. | - | - | borderline |
| Others | Passed away during<br>early childhood (ages 0-5),<br>Necrotizing enterocolitis,<br>Unspecified Hepatic<br>Dysfunction,<br>Conjugated<br>hyperbilirubinemia,<br>Thrombocytopenia,<br>Seizures. | Passed away during<br>early childhood (ages 0-5),<br>Distended bowel,<br>Necrotizing enterocolitis,<br>Thrombocytopenia. | Passed away during<br>early childhood (ages 0-5),<br>Distended bowel,<br>Unspecified Hepatic<br>dysfunction,<br>Conjugated<br>hyperbilirubinemia,<br>Thrombocytopenia,<br>Postmortem liver biopsy<br>showed severe<br>cholestasis and ductular<br>proliferation with<br>significant portal fibrosis. | Prominent glabella,<br>Forehead hemangioma,<br>Open mouth, Mandible<br>ankylosis, ankyloglossia. | Generalized hypotonia,<br>Nissen fundoplication for<br>gastro-esophageal reflux<br>at age 0-5,<br>Hypo immunoglobulins,<br>Protein losing gastro-<br>enteropathy,<br>Hypertrichosis on the<br>back. |

\*, Age at the time of passing in days/months; -, not present; +, present; BW, body weight; HC, head circumference; SD, standard deviation; ASD, atrial septal defect; VSD, ventricular septal defect; n.a., information not available.

**Supplementary Table 2.**
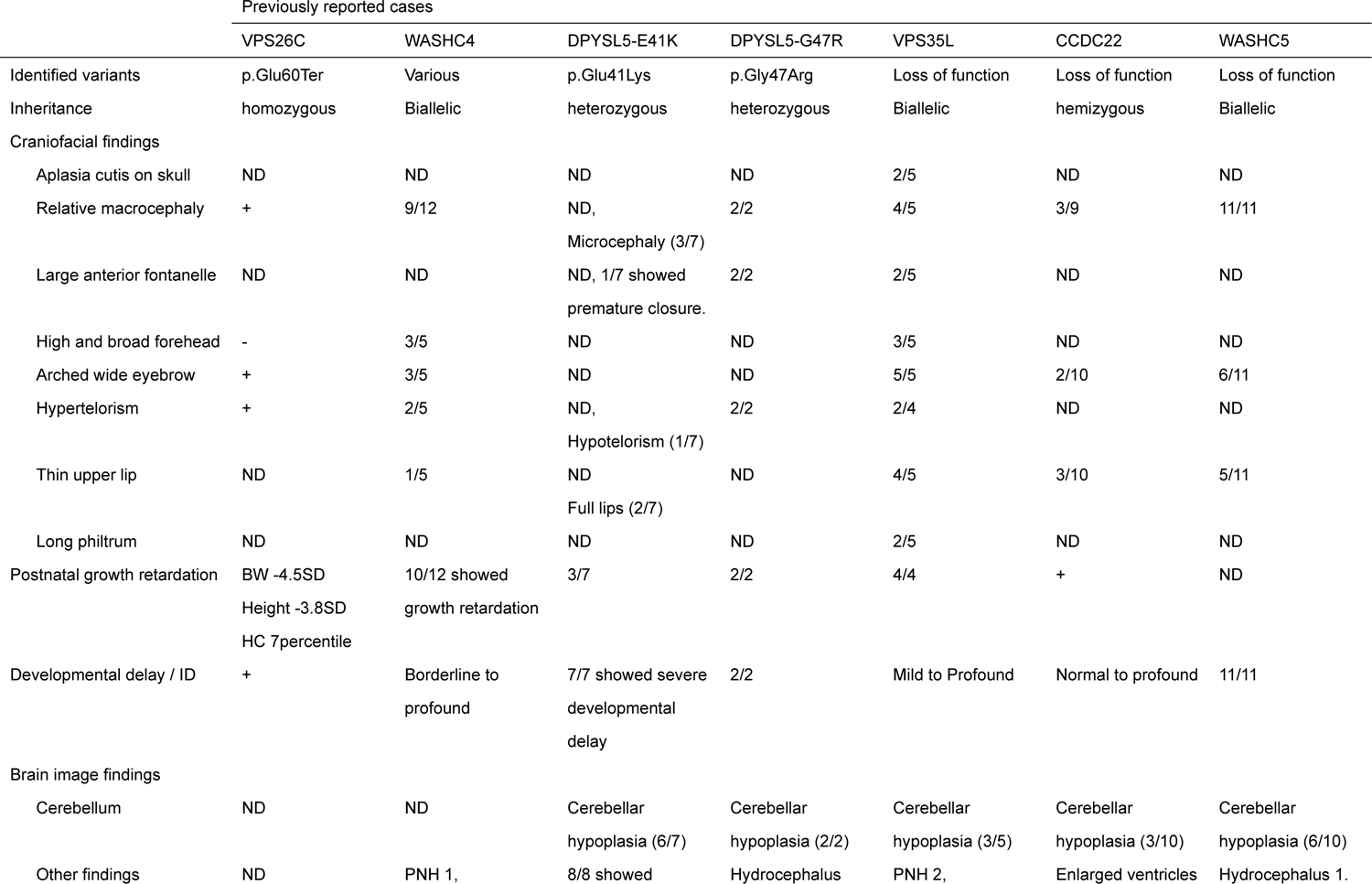
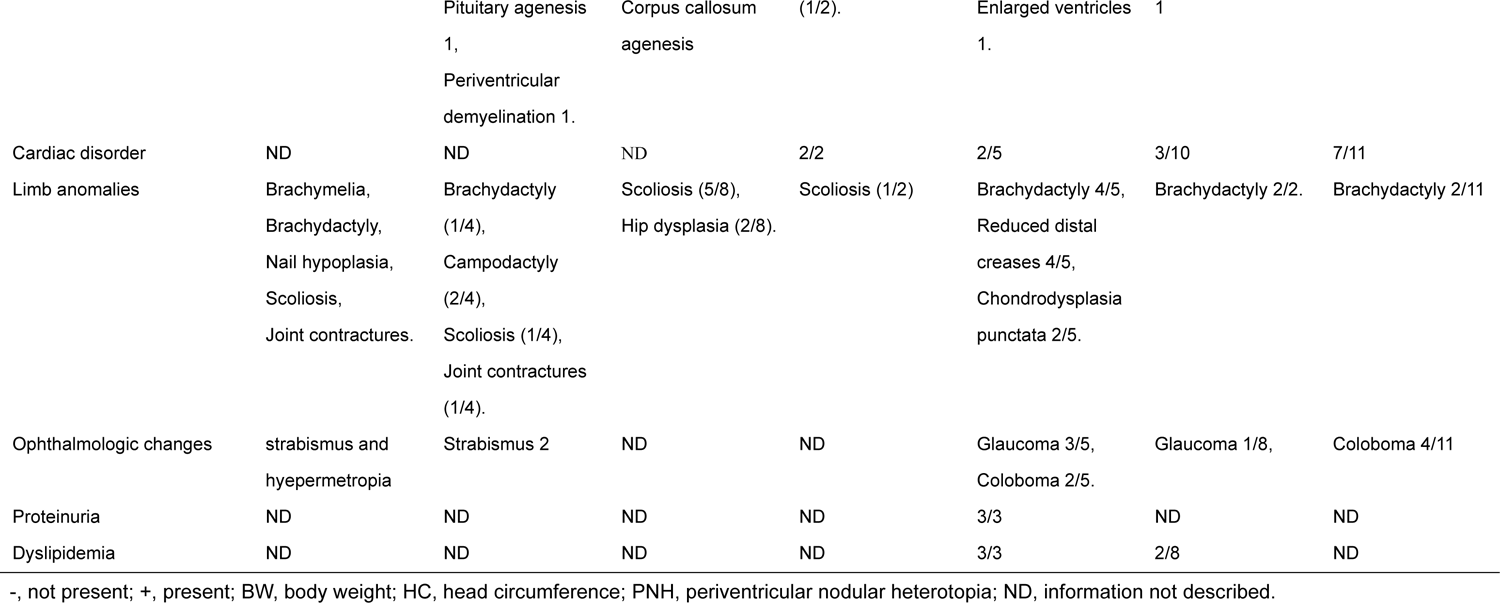
Clinical comparison among patients with 3C-Ritscher Schinzel syndrome and other Commander/WASH associated gene mutations.

